# Characterising information gains and losses when collecting multiple epidemic model outputs

**DOI:** 10.1101/2023.07.05.23292245

**Authors:** Katharine Sherratt, Ajitesh Srivastava, Kylie Ainslie, David E. Singh, Aymar Cublier, Maria Cristina Marinescu, Jesus Carretero, Alberto Cascajo Garcia, Nicolas Franco, Lander Willem, Steven Abrams, Christel Faes, Philippe Beutels, Niel Hens, Sebastian Müller, Billy Charlton, Ricardo Ewert, Sydney Paltra, Christian Rakow, Jakob Rehmann, Tim Conrad, Christof Schütte, Kai Nagel, Sam Abbott, Rok Grah, Rene Niehus, Bastian Prasse, Frank Sandmann, Sebastian Funk

## Abstract

**Background:** Collaborative comparisons and combinations of epidemic models are used as policy-relevant evidence during epidemic outbreaks. In the process of collecting multiple model projections, such collaborations may gain or lose relevant information. Typically, modellers contribute a probabilistic summary at each time-step. We compared this to directly collecting simulated trajectories. We aimed to explore information on key epidemic quantities; ensemble uncertainty; and performance against data, investigating potential to continuously gain information from a single cross-sectional collection of model results.

**Methods:** We compared July 2022 projections from the European COVID-19 Scenario Modelling Hub. Five modelling teams projected incidence in Belgium, the Netherlands, and Spain. We compared projections by incidence, peaks, and cumulative totals. We created a probabilistic ensemble drawn from all trajectories, and compared to ensembles from a median across each model’s quantiles, or a linear opinion pool. We measured the predictive accuracy of individual trajectories against observations, using this in a weighted ensemble. We repeated this sequentially against increasing weeks of observed data. We evaluated these ensembles to reflect performance with varying observed data.

**Results:** By collecting modelled trajectories, we showed policy-relevant epidemic characteristics. Trajectories contained a right-skewed distribution well represented by an ensemble of trajectories or a linear opinion pool, but not models’ quantile intervals. Ensembles weighted by performance typically retained the range of plausible incidence over time, and in some cases narrowed this by excluding some epidemic shapes.

**Conclusions:** We observed several information gains from collecting modelled trajectories rather than quantile distributions, including potential for continuously updated information from a single model collection. The value of information gains and losses may vary with each collaborative effort’s aims, depending on the needs of projection users. Understanding the differing information potential of methods to collect model projections can support the accuracy, sustainability, and communication of collaborative infectious disease modelling efforts.

**Data availability:** All code and data available on Github: https://github.com/covid19-forecast-hub-europe/aggregation-info-loss

## Background

During outbreaks of infectious disease, it is critical to account for the uncertainty of future disease incidence in order for public health decision-makers to fully evaluate risk [1], [2]. Infectious disease modellers use a variety of approaches to meet this demand for information. A common challenge is the representation of multiple sources of uncertainty, both within each model as well as across separate model projections [3], [4]. In recognising this challenge, infectious disease modelling has seen an increasing emphasis on both probabilistic modelling methods, together with collaborative approaches to modelling [5], [6].

Probabilistic infectious disease models can address the challenge of uncertainty by simulating the complex and changing real-world process of disease transmission. Modellers must handle stochasticity in transmission dynamics, often using observed data to estimate model parameters and latent trajectories that are themselves uncertain. Each such model can generate any number of simulated trajectories, and modellers choose at what point to conclude there are sufficient iterations to reach a stable distribution of possible outcomes. The output of these simulations can then be summarised to calculate quantities of interest, such as weekly incidence of infections or cases.

When creating models to characterise the future, modellers have often drawn a distinction in the meaning of uncertainty between forecast compared to scenario projections [7]. Forecasts are predictions of future epidemic trajectories, and the probabilities assigned to different outcomes quantify the belief of the forecaster that these may or may not happen. In addition to potential fundamental limits to predictability, forecasts are usually reliable for, at best, a few generations of transmission [8] because of unmodelled factors affecting future transmission such as behavioural or policy changes, heterogeneity in transmission risk, or the emergence of new variants of different transmissibility or severity.

In contrast, scenarios are projections attuned to a particular context by being conditioned on specific factors whose futures may not be quantitatively predictable, such as options for policy interventions [9], [10]. Probabilities of future outcomes as stated by scenario models should be interpreted as valid only under the specific circumstances given by the scenario but not otherwise, without specifying any probability of the scenario itself occurring. Because of this difference, forecasts can be evaluated by confronting them with future data as it becomes available, while this evaluation is more challenging for scenarios where predictive performance will always depend on a combination of adequacy of the chosen assumptions (e.g. on pathogen biology, human behaviour and government policy), with adequacy of the model in reflecting these assumptions.

Infectious disease modelling collaborations aim to bring together models that project the future using diverse methods [6]. Each collaboration sets a clearly defined target for projections, communicates this target to multiple independent modellers, and collects model results in a standardised format. This standardisation allows for a like-for-like comparison of varying modelling methods’ results and accompanying uncertainty. Ensemble methods can then combine results across models. Typically, this creates a more comprehensive and robust projection [11] or reflection of expert judgement [12].

Formal, large-scale modelling collaborations have, so far, been used for influenza, Ebola, Zika, dengue fever, and COVID-19 [6]. In the case of COVID-19, a number of policy-facing research groups have set up collaborations to collate forecasts and scenarios [13], [14], [15], [16], and there is a substantial effort towards expanding the practice of ensemble projections of infectious disease spread and burden. Ongoing work evaluating these efforts has focused on assessing the output of past and current ensemble modelling projects. This has included evaluating differing performance among individual models [17], [18], [19], and a variety of methods for creating ensembles from multiple models [11], [16], [20], [21].

The standardised format in which model projections are collected is key to meeting such projects’ aims of comparing information from multiple models. The most common approach to this is to collect descriptive statistics from each model at each given time step. In this format, each modeller submits values across a pre-specified set of quantiles in order to represent uncertainty in their projection. The benefits of this system include that it should accurately represent an underlying distribution of outcomes while being storage-efficient [22], it is not restricted to probabilistic models producing simulations, and it allows for quantitative evaluation against observed data [5]. Various methods for subsequent combination from quantiles depend on the view taken of uncertainty between and across model projections [21].

However, using quantile intervals separately across each time step may lose information pertinent to epidemic decision making. As a quantile representation provides a summary across trajectories at each time step, it has no theoretical continuity through the time-series. This does not permit aggregation over time to calculate cumulative totals or means, and may misrepresent time-series characteristics including epidemic peak size or timing [23]. Whilst some of these can be remedied by also collating quantiles of cumulative quantities, these still lose some of the temporal information contained in the full joint probability distribution across all future time points.

An alternative method of collecting output from multiple probabilistic models is to collect the individual simulated trajectories produced by each modeller. Each simulated trajectory comprises a single value for each time step, with modellers contributing some number of these trajectories. Each trajectory retains its own time-series characteristics, and these can therefore be summarised across different models. Collecting trajectories also creates potential for the analysis and combination of each trajectory independently from the originating model’s total output. One option could include comparing each trajectory to observed data as it becomes available, even after the time of collecting model outputs. This would enable creating an ensemble projection that is conditioned on the observed accuracy of each individual trajectory. As further observed data become available, this ensemble could be updated to create a single combined projection that continuously reflects the changing performance of each trajectory. This would act similarly to methods of particle filtering in continuously conditioning on past behaviour.

We aim to explore aspects of information gains and losses from these two methods of collecting multiple model results. We contrast collecting a set of simulated trajectories, against collecting a summary at quantile intervals of those trajectories. We use the setting of the European COVID-19 Scenario Hub, where the use of quantile summaries was replaced in mid-2022 by collecting trajectories. These trajectories represent random samples from the collection of all possible trajectories of each model consistent with a given scenario and the data available up to the time at which the simulation was generated.

In this work, we assess the impact of the collection method when seeking information about policy-relevant epidemic characteristics, including cumulative totals, timing of peaks, and the extent of uncertainty across multiple models. We then explore the information gained by the ability to compare modelled epidemic trajectories to observed data as this becomes available over time. We use this to create a multi-model ensemble which weights across all available trajectories by their past accuracy. This demonstrates the potential to continuously gain information from only a single cross-sectional collection of model results. Understanding the potential sources of information gains and losses when collecting multiple model projections may support improving the accuracy, reliability, and communication of collaborative infectious disease modelling efforts.

## Methods

### Study setting

In this work we use projections from Round 2 of the European COVID-19 Scenario Modelling Hub [20]. The European COVID-19 Scenario Hub was launched in March 2022 to reflect demand for the ECDC to support longer term European policy planning. It used the existing US Scenario Hub [15] as a basis for Hub infrastructure and methods. Modelling teams were recruited by word of mouth to join a series of collaborative workshops, approximately fortnightly from March through June 2022. In these sessions both policy-focussed colleagues from the ECDC and modelling-focussed researchers co-developed a set of four scenarios. Each scenario represented a combination of two possible epidemiological and policy changes that could impact the incidence of COVID-19 across Europe in the medium term.

Teams were asked to project the incidence of COVID-19 infections, cases, deaths, and hospitalisations in 32 European countries over the next year. To facilitate comparison across models, we identified and agreed a common set of key assumptions and parameters to be used by all models in each scenario as well as standard data sets to which to compare the model outputs where available. Modellers uploaded projections to a Github repository, and we summarised results across models, with a focus on targets with three or more different models. Over 2022 this process was repeated four times to explore a variety of different scenarios. In total nine separate teams submitted projections, with six teams contributing to each round.

Over June 2022 (Round 2), we specified four scenarios (A-D) as: an autumn second booster campaign among the population aged over 60 (scenarios A/C), or over 18 (scenarios B/D); and future vaccine effectiveness as ‘optimistic’ (equivalent to the effectiveness as of a booster vaccine against the Delta SARS-CoV-2 variant; scenarios A/B); or ‘pessimistic’ (as against variants Omicron BA.4/BA.5/BA.2.75; scenarios C/D). Modellers were asked to start their projections from 24th July 2022, meaning that even if data were available beyond this date they were not to inform calibration of the model. Modellers were asked to submit up to 100 simulations, each reflecting a trajectory of weekly incidence of reported cases and deaths over time for a given projection target. Modellers were informed that data presented on the Johns Hopkins University dashboard was to be used for future comparison to data [24]. In practice some of the models were not calibrated to reported cases and therefore used symptomatic cases as a proxy (see model details in Supplement). Simulations were to represent random samples from the distribution of simulation trajectories consistent with the given scenario that each modelling team produced. We have published full scenario details including shared parameters, all teams’ projections, and summary results online [25].

This work specifically focuses on contrasting the sampled simulated trajectories with their representation in time-specific quantiles. We collected raw data in the form of up to 100 trajectories from each model for each projection target. We used these data to retrospectively create a marginal fixed-time quantile representation of results from each model and target. Following the current submission procedure across COVID-19 Modelling Hubs for an individual model, we calculated a median and 22 further quantiles for each week using the values of the trajectories in that week, separately for each scenario. We processed all data in R with code available online [26].

### Characterising potential information gains and losses

First we considered information about key epidemic characteristics. At the time the projections were in production, discussion with the ECDC modelling team led to an interest in: estimates of incidence over time; cumulative values over different periods; and number of distinct peaks, size, and timing of peak incidence over the projection period.

When projections were available, we estimated these characteristics from the simulated trajectories. We summed incidence over time to produce a cumulative total from each trajectory. We assessed the size of the expected burden of each target relative to a known threshold by comparing the cumulative projected total to the cumulative total of the preceding year. We identified peaks in each simulated trajectory as the local maxima in a sliding window of five weeks, using the ggpmisc R package [27]. We chose a sliding window of five weeks to capture each distinct peak while avoiding detecting noise in each trajectory. We summarised across the individual peaks detected in each trajectory using quantiles at each weekly time-step, to produce a range indicating possible peak timing and maximum values across all trajectories. We produced a real-time report of this summary at the time that projections became available in July 2022.

In further retrospective analysis, we compared the use of a standard unweighted ensemble to express uncertainty across multiple models in the two representations. We created an ensemble projection from first combining all individual simulated trajectories with equal weight for each scenario, location, and outcome target. Next, we took model-specific quantiles from each model’s distribution of trajectories at each time point, for each scenario, location, and outcome target. We used each set of quantiles to create linear opinion pool ensembles (LOP), which use linear extrapolation between the given quantiles to estimate the cumulative distribution function in order to then randomly sample trajectories to aggregate, again with equal weight; and a quantile-average ensemble, which takes the median across the different models’ values at each quantile and time step. The LOP and quantile-average ensembles have both been used to produce ensemble projections across multiple epidemiological forecasts [21], [11], [16]. To assess the difference in uncertainty across the two ensembles, we compared the mean of the values at each quantile across all time points, outcomes and scenarios.

Lastly, we evaluated the performance of each simulated trajectory against proximity to observed data, and used this to weight an ensemble of trajectories (as above). To measure performance, we calculated the mean absolute error (MAE) for each trajectory, where the MAE is the average of the difference from observed data across all available time points for a single projection. We created a weighted ensemble from all trajectories for a country (not further separating by scenario or model) using the inverse MAE for each trajectory as a weight. To calculate weighted quantiles we used a Harrel Davis weighted estimator [28] from the *cNORM* R package (v3.0.2) [29]. As above, we calculated 23 quantiles including the median to express uncertainty.

We repeated this process to create a sequence of ensembles with changing weights over time. We created the first weighted ensemble after 4 weeks of observed data, and then created consecutive ensembles with weights re-calculated weekly to use up to the maximum available 29 weeks of observed data (to 11 March 2023). This showed varying lengths of projections repeatedly conditioned on simulated trajectories’ performance against increasing data over time.

We evaluated the predictive performance of these sequences of weighted ensembles. We transformed forecasts and observed data to a logarithmic scale, as this allows a more consistent evaluation across varying magnitudes and better reflects the exponential nature of epidemic processes [30]. We then calculated the weighted interval score for each forecast, as a quantitative performance measure that evaluates across both the accuracy and the dispersion of probabilistic forecasts [5]. In the same way we evaluated the unweighted ensemble of trajectories described above, and used this as a relative baseline with which to compare the effect of weighting individual trajectories on ensemble performance.

## Results

A total of six modelling teams contributed projections for various targets to the European COVID-19 Scenario Hub in Round 2. Here we focus on multi-model comparison and include only projection targets with three contributing models. These targets included 52 weeks’ case and death incidence for the Netherlands and Belgium, and 41 weeks’ case incidence for Spain.

Five teams contributed projections for these targets. Three teams used compartmental models, one an agent-based model, and one a machine learning method (see Supplement). Four models generated 100 simulated trajectories, and one 96 trajectories (implying a slightly smaller weight to this model in trajectory-based aggregates). In total, we consider 294,816 data points from 5,920 trajectories, where each data point is the estimated weekly incidence in a simulated trajectory of an outcome in a target country and scenario over up to one year (figure 1.i).

**Figure 1.**
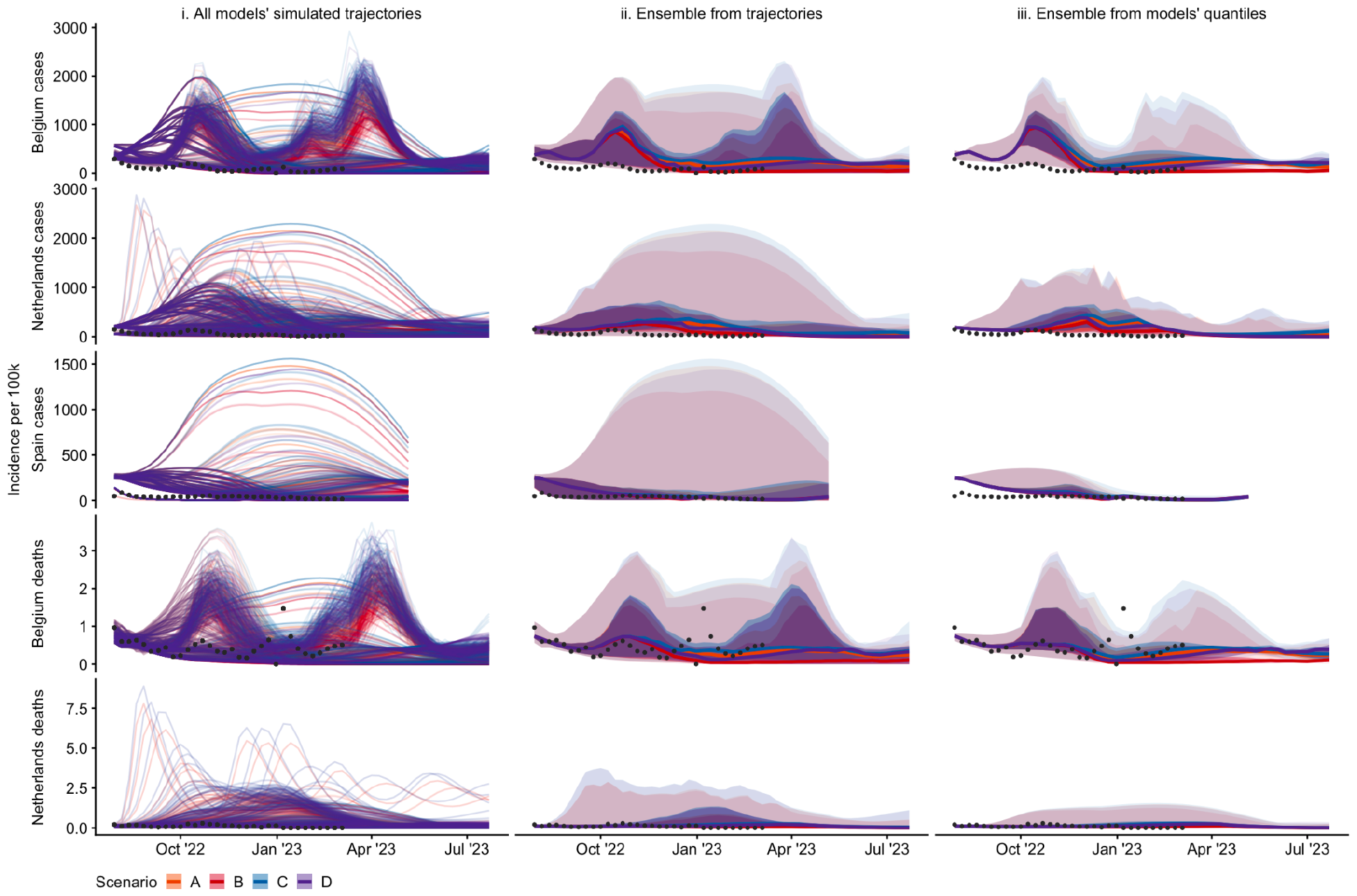
Projections of incidence per 100,000 population, by country (row) and aggregation method (column) showing median, 50%, and 99% probabilistic intervals (increasingly shaded ribbons), for each scenario, using: i) no ensemble method (100 simulated trajectories per model, or 96 in case of one of the models); ii) quantile intervals of the distribution across all simulated trajectories; ii; a median across each model’s projections at a given quantile interval. We do not show the linear opinion pool ensemble here as results are near-identical to the ensemble drawn directly from trajectories (ii)). Scenarios included: an autumn second booster vaccine campaign among population aged 18+ (scenarios B & D) or 60+ (scenarios A & C); where vaccine effectiveness is ‘optimistic’ (effectiveness as of a booster vaccine against Delta; scenarios A & B) or ‘pessimistic’ (as against BA.4/BA.5/BA.2.75; scenarios C & D). See Supplement for further detail on individual models’ trajectories.

Aggregating across simulated trajectories from multiple models allowed access to information about various epidemic characteristics. These included cumulative totals, and peak size and timings (see contemporaneous report reproduced in the Supplement). By summarising across the peaks of each individual trajectory, we were able to create an estimate of uncertainty around the size and timing of peaks for each target. We were also able to summarise cumulative outcomes. For example, across all 5920 trajectories for all targets and scenarios, 10% saw a cumulative total exceeding the preceding year. These epidemic characteristics could not be meaningfully estimated from the same results summarised into quantiles.

We compared information loss in the aggregation of simulated trajectories into ensemble projections (figure 1). We compared an ensemble taken from all trajectories (figure 1.ii) with a linear opinion pool (not shown), and the quantile-average ensemble (figure 1.iii). We noted that a linear opinion pool ensemble produces near-identical results to taking an ensemble directly from trajectories. Across all projection targets, we observed substantially increased uncertainty in an ensemble that aggregated either directly from trajectories, or via linear opinion pool, compared to a quantile-average ensemble. This represented the wider variety of epidemic shapes projected by different models. For example, the credible interval of projections for Spain included high autumn-winter incidence, while for Belgium gave greater credibility to multiple peaks of incidence. These were not observed in the interval projections of an ensemble derived from models’ quantiles.

We quantified the range of uncertainty between each ensemble by comparing the mean of values at each quantile across all time points and scenarios (supplementary figure 1A). All ensembles produced similar values around the centre of the distribution, with no noticeable difference between the median values of each projection. However, across all five targets we observed that an ensemble based on either simulated trajectories, or an LOP ensemble, produced sharply increasing uncertainty between the 90% to 98% intervals. For example, at the upper 98% probability interval, ensemble projections for cases in Spain averaged nearly six times higher incidence when drawn directly from trajectories compared to when drawn from a median of three models’ quantiles (respectively averaging 1016 and 173 weekly new cases per 100,000 population).

We then considered an ensemble of individual trajectories each weighted against a sequentially increasing amount of observed data (figure 2). We note that models used a variety of methods and may have been calibrated to alternative data sources (see Supplement). In comparison to the unweighted ensemble (shown in grey), we observed reduced uncertainty across weighted ensemble projections. Compared to conditioning on data up to 16 weeks before, adding 8 weeks of additional data in weighting case projections reduced the upper 98% bound of uncertainty by at least 5% and up to 30% on average (supplementary figure 1B). The accuracy-weighted contribution of each trajectory to an ensemble varied substantially between models and targets, and over time. For example, in Spain each trajectory’s weight remained stable after mid December 2022, reflecting the data by effectively downweighting those trajectories projecting sustained high incidence over winter (see figure 1i).

**Figure 2.**
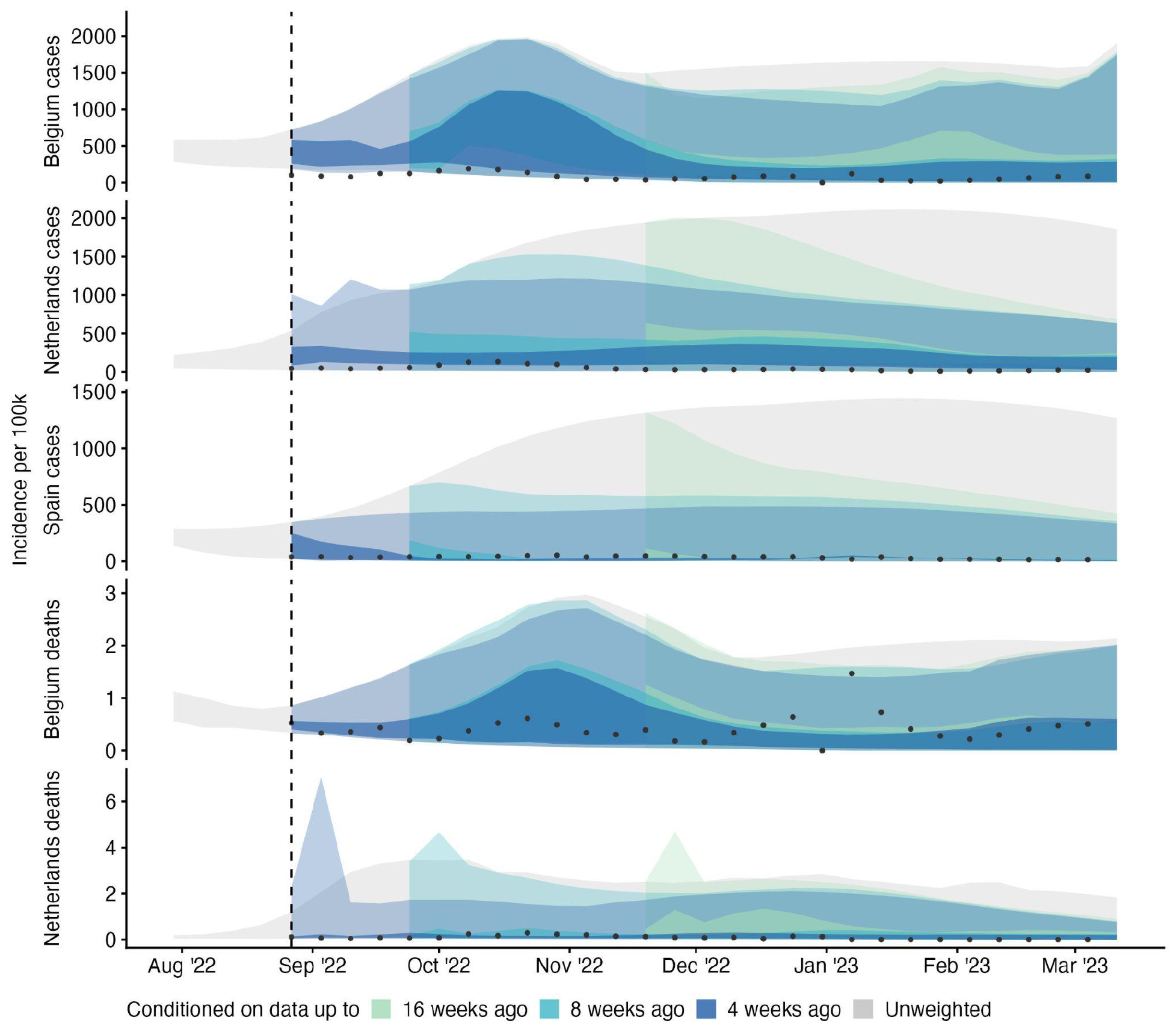
Ensemble forecasts of incidence by target, using no weighting (grey ribbon), or 4, 8, and 16 weeks ahead of available data, with available data increasing weekly over time (coloured ribbons); showing 50% and 99% credible intervals. Each simulated trajectory started from 30 July 2022 and was weighted using its inverse mean absolute error against available data. We used at least 4 and up to 31 weeks of this observed accuracy data.

We used this information to create consecutive weekly ensembles, with weights updating as increasing observed data became available to measure trajectories’ accuracies. In the combined (weighted interval) score, forecasts using weighted trajectories generally performed similarly to the unweighted equivalent, with a median relative WIS among the weighted ensembles of 0.99 (IQR: 0.89-1.05; supplementary figure 2).

When using the full 31 weeks of available data, a weighted ensemble performance improved compared to projections made without weighting on accuracy (with a median relative WIS across targets of 0.77 compared to the baseline of 1). However, this improvement was not linearly correlated with increasing data, and the relationship varied by target (figure 3). Weighted forecasts that used only a few data points of trajectories’ accuracy performed similarly or poorly compared to the unweighted ensemble.

**Figure 3.**
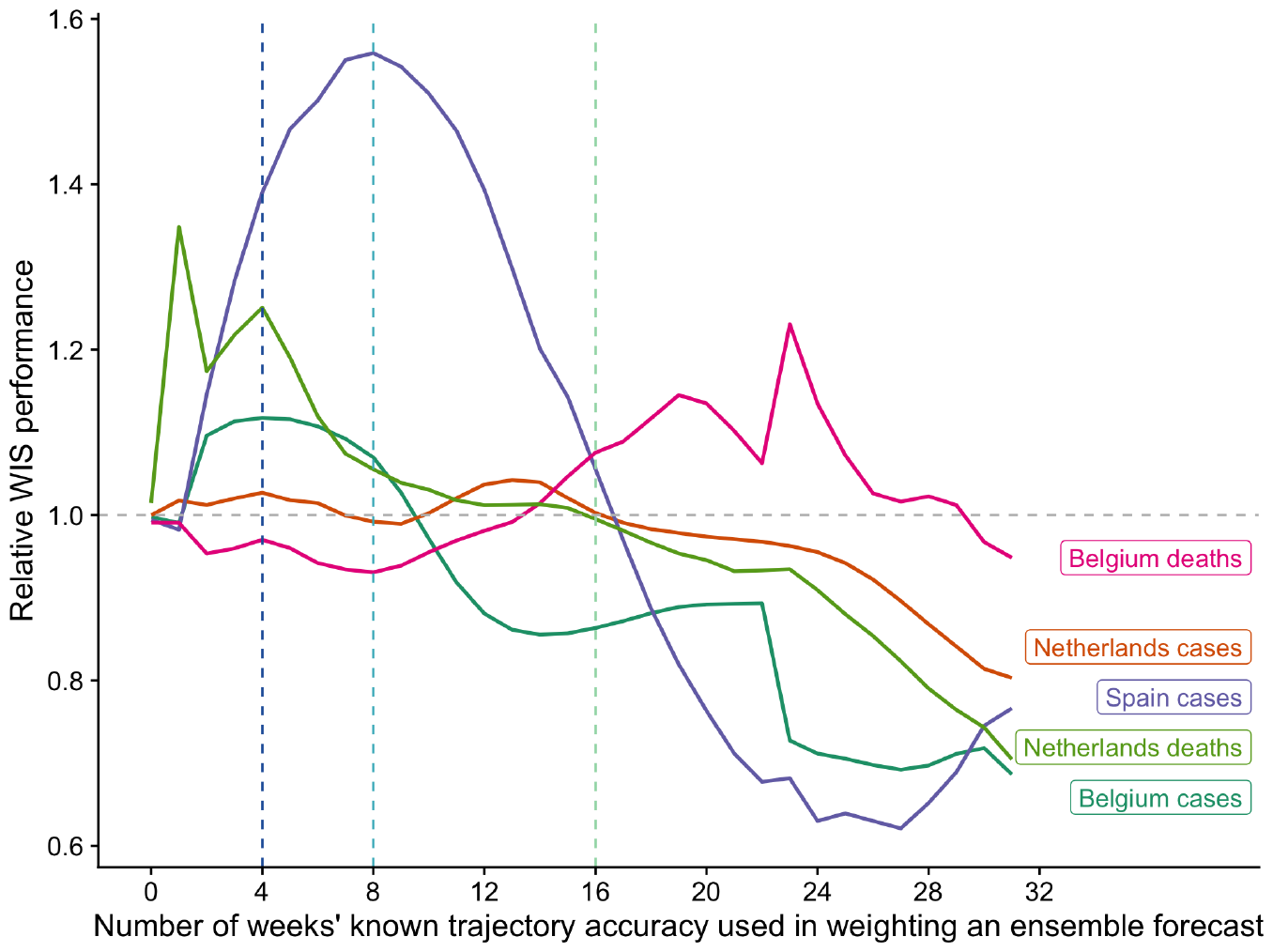
Predictive performance of weighted ensembles by projection target. Weighted ensembles were created using a weighted median, where the weight of each trajectory was determined by its previous accuracy in predicting between 0 and 31 weeks of observed data (x axis). The performance of each ensemble is measured by the weighted interval score (WIS); a lower WIS score indicates better performance of the weighted ensemble than the simple unweighted median ensemble of all trajectories (reference line at 1).

However, among three targets, using more accuracy data gave a stable or consistently improved performance (after 10 weeks for cases in Belgium and 17 weeks for cases and deaths in the Netherlands). This was also true of cases in Spain, with worse performance compared to the unweighted ensemble when using up to 9 weeks of data, and improving and then better relative performance after 17 weeks of data, until performance worsened once more after 27 weeks of accuracy data. In contrast, forecasts of deaths in Belgium were better with fewer weeks of accuracy data, and weighting with between 14-26 weeks’ data produced a worse performance than the unweighted ensemble of trajectories.

## Discussion

A significant part of the value of collaborative infectious disease modelling projects comes from the standardisation of model output across varying numbers of model teams, methods, and simulations. We compared two methods of collecting information from multiple models’ projections of an epidemic. We took three scenario models for each of five projection targets, and contrasted collecting a sample of up to 100 simulated trajectories against collecting quantile intervals of those trajectories at each time step.

We found that collecting simulated trajectories enabled analysis of trajectory shapes, peaks, and cumulative total burden. We observed that trajectories contained a right-skewed probabilistic distribution, which meant that ensembles either directly from trajectories, or using a linear opinion pool method, increasingly diverged from the quantile-average ensemble in projecting the outer upper limit of the probabilistic distribution. We also found that collecting trajectories could be used to create a competitively performing ensemble based on continuous predictive performance.

The common practice of collecting a standardised set of quantile intervals has several advantages. Firstly, combining across a set of quantiles should accurately represent the underlying distribution [22], and we observed that the linear opinion pool (based on a combination of quantiles) produced a near-identical ensemble as that created directly from combining individual trajectories. This suggests that the LOP ensemble may be the best choice for reflecting the widest range of uncertainty in settings where model results are only collected in quantiles [21], while noting that in order to create a LOP ensemble quantiles of cumulative rather than incident quantities need to be collected. Furthermore, our results suggest little information about uncertainty is lost when using quantile outputs to compare the central estimates from different models. This is a useful validation for collecting multiple model results in any format when the purpose is short-term situational awareness.

Further advantages include where collecting quantile outputs also allows for a broader range of modelling methods, including quantile regression, that directly create quantile outputs rather than a joint distribution over time. Additionally, a single set of quantiles can be held in comma-separated value (csv) files of easily manageable size, requiring minimal technical knowledge of big data storage solutions or processing. This has been important in the past given a lack of readily available skills or investment in software for emergency outbreak settings. However this argument weakens with mounting evidence that this type of under-resourcing hampers outbreak response [31].

An alternative method for multiple model collection is directly collecting models’ trajectories, with the advantage of retaining each trajectory’s time-dependence. We observed greater availability and flexibility of accessing information from this method in contrast to collecting quantile distributions. This was evident when comparing the tails of multiple distributions in a quantile-average ensemble, assessing the number of projected waves or the risk of crossing a specific threshold such as the burden in the preceding year, or in reevaluating projections against reported data. These analyses could also be conducted after collecting model outputs, making the method of collecting trajectories more flexible to the needs of one or multiple end-users. In particular these areas of information are more likely to be relevant to longer term preparedness and mitigation. As a result, we suggest the impact of information gains and losses from model collection may differ depending on the aim of a multi-model comparison.

Our findings comparing quantile with trajectory model outputs are compatible with ongoing work addressing issues from the loss of epidemic shape. From point forecasts, recent forecasting work has created an ensemble from multiple point forecasts in terms of similarity to canonical curve shapes [32]. From probabilistic models it is also possible to create an ensemble of many trajectories using the centrality of each curve as a weight in a curve boxplot [23].

We have also demonstrated the potential for unique information gains when collecting simulated trajectories by assessing their performance against observed data. By conditioning the weight of each trajectory in an ensemble on subsequently observed data, we were able to create an ensemble that excluded entire trajectories, or epidemic curves, based on dependence to unrealised events. This typically either matched or reduced the uncertainty of an unweighted equivalent ensemble, and in some settings performed better overall than the unweighted equivalent.

This suggests an additional way in which collaborative modelling efforts can respond to changing outbreak dynamics and policy needs. For our setting, model results had originally been created based on a set of four scenarios relevant to policy decisions to be made in spring/summer 2022. However, given the complex dynamics of disease transmission, no predefined future scenario is likely to accurately predict eventual reality. Among four scenarios with deliberately contrasting assumptions, most of these assumptions will be disproven by observation over time. Meanwhile, when scenario modelling outputs are collected as quantiles at each time-point, they lose their time-dependence and thus cannot be interpreted except in the light of an increasingly obsolete scenario context.

By focussing on individual model simulations in this work, we were able to abstract away from the context in which model results were created. We weighed each trajectory using only its past accuracy against observed data, regardless of the modelling technique, original scenario, or parameter values from which it arose. From this we created an ensemble that did not reflect any particular scenario assumptions, but only the time-varying accuracy of each trajectory. This meant we were able to continue to use trajectories in an ongoing evaluation, increasing the useful life of the results from a single cross-sectional collection of multiple model output.

This could be particularly useful when repeated rounds of model collection are time-intensive or computationally expensive, such as for individual-based models, or where personnel resources are constrained such as in an ongoing outbreak with potentially many competing priorities. Whilst beyond the scope of this work, future work in this area could also investigate model weights in order to rank trajectories from each scenario by proximity to observed trajectories and potentially interpret this as proximity of the given scenario assumptions to reality.

We highlight several important limitations to our comparison of information gains and losses between methods of collecting model output. In the first part of this work, we represent an analysis of trajectories that reflects our work in real-time response to the needs of policy decision-making. We did not consider alternative approaches to time-series analysis that would likely make the analysis of trajectories more robust, for example in calculations of peaks or wave durations.

In our comparison to quantile distributions, our method of collecting simulated trajectories was not specifically designed for this comparative purpose, and as a result our findings are difficult to interpret. In this work we did not attempt to characterise how many samples might be sufficient to appropriately represent a probabilistic distribution in comparison to a quantile representation. For example, in some cases the collated trajectories were already subsampled from model runs conducted by individual teams. In contrast, in a situation with low sampling sizes of trajectories from each model, a quantile representation might provide a more stable representation.

We suggest that further work should characterise and standardise sampling techniques for model simulations in multi-model comparisons. Future study designs could focus on collating multiple representations (e.g. time-sliced quantiles and trajectories) from contributing teams directly for comparison, or collate arbitrary numbers of feasible simulated trajectories and re-weigh according to the number of simulations. Our work also demonstrates the importance of investing in and developing capacity to store and use simulation outputs rather than fixed-time quantile probabilities for well founded intercomparison modelling projects.

To conclude, we observed several information gains from collecting modelled trajectories rather than summarised quantile distributions. We highlight the potential to create continuous new information from a single collection of model output. Working from combined simulations offers the opportunity to explore creating ensembles by the shape of epidemic curve that can be updated over time, and for more detailed quantitative evaluations against observed data, such as in projected peaks or cumulative totals. We believe our findings apply whether projections are conditioned on the context of the present (as in forecasts), or on schematic futures (as in scenarios). However, the value of different information gains and losses may vary with the aims of each collaborative effort, depending on the requirements and flexibility required by projection users. Understanding potential information gains and losses when collecting model projections can support the accuracy, reliability, sustainability, and communication of collaborative infectious disease modelling efforts.

## Data Availability

All code and data available on Github: https://github.com/epiforecasts/aggregation-info-loss

## References

[1] J. Zelner, J. Riou, R. Etzioni, and A. Gelman, ‘Accounting for uncertainty during a pandemic’, Patterns, vol. 2, no. 8, Aug. 2021, doi: 10.1016/j.patter.2021.100310.

[2] S.-L. Li et al., ‘Essential information: Uncertainty and optimal control of Ebola outbreaks’, Proc. Natl. Acad. Sci., vol. 114, no. 22, pp. 5659–5664, May 2017, doi: 10.1073/pnas.1617482114.

[3] R. McCabe et al., ‘Communicating uncertainty in epidemic models’, Epidemics, vol. 37, p. 100520, Dec. 2021, doi: 10.1016/j.epidem.2021.100520.

[4] B. Swallow et al., ‘Challenges in estimation, uncertainty quantification and elicitation for pandemic modelling’, Epidemics, vol. 38, p. 100547, Mar. 2022, doi: 10.1016/j.epidem.2022.100547.

[5] J. Bracher, E. L. Ray, T. Gneiting, and N. G. Reich, ‘Evaluating epidemic forecasts in an interval format’, PLOS Comput. Biol., vol. 17, no. 2, p. e1008618, Feb. 2021, doi: 10.1371/journal.pcbi.1008618.

[6] N. G. Reich et al., ‘Collaborative Hubs: Making the Most of Predictive Epidemic Modeling’, Am. J. Public Health, vol. 112, no. 6, pp. 839–842, Jun. 2022, doi: 10.2105/AJPH.2022.306831.

[7] M. Lipsitch, L. Finelli, R. T. Heffernan, G. M. Leung, and S. C. Redd; for the 2009 H1N1 Surveillance Group, ‘Improving the Evidence Base for Decision Making During a Pandemic: The Example of 2009 Influenza A/H1N1’, Biosecurity Bioterrorism Biodefense Strategy Pract. Sci., vol. 9, no. 2, pp. 89–115, Jun. 2011, doi: 10.1089/bsp.2011.0007.

[8] K. Sherratt et al., ‘Predictive performance of multi-model ensemble forecasts of COVID-19 across European nations’, eLife, vol. 12, p. e81916, Apr. 2023, doi: 10.7554/eLife.81916.

[9] M. C. Runge et al., ‘Scenario Design for Infectious Disease Projections: Integrating Concepts from Decision Analysis and Experimental Design’. medRxiv, p. 2023.10.11.23296887, Oct. 12, 2023. doi: 10.1101/2023.10.11.23296887.

[10] T. Rhodes, K. Lancaster, S. Lees, and M. Parker, ‘Modelling the pandemic: attuning models to their contexts’, BMJ Glob. Health, vol. 5, no. 6, p. e002914, Jun. 2020, doi: 10.1136/bmjgh-2020-002914.

[11] E. L. Ray et al., ‘Ensemble Forecasts of Coronavirus Disease 2019 (COVID-19) in the U.S.’, medRxiv, p. 2020.08.19.20177493, Aug. 2020, doi: 10.1101/2020.08.19.20177493.

[12] K. Shea et al., ‘Harnessing multiple models for outbreak management’, Science, vol. 368, no. 6491, pp. 577–579, May 2020, doi: 10.1126/science.abb9934.

[13] S. Funk et al., ‘Short-term forecasts to inform the response to the Covid-19 epidemic in the UK’, medRxiv, p. 2020.11.11.20220962, Nov. 2020, doi: 10.1101/2020.11.11.20220962.

[14] E. Y. Cramer et al., ‘The United States COVID-19 Forecast Hub dataset’. medRxiv, p. 2021.11.04.21265886, Nov. 04, 2021. doi: 10.1101/2021.11.04.21265886.

[15] R. K. Borchering, ‘Modeling of Future COVID-19 Cases, Hospitalizations, and Deaths, by Vaccination Rates and Nonpharmaceutical Intervention Scenarios — United States, April–September 2021’, MMWR Morb. Mortal. Wkly. Rep., vol. 70, 2021, doi: 10.15585/mmwr.mm7019e3.

[16] K. Sherratt et al., ‘Predictive performance of multi-model ensemble forecasts of COVID-19 across European nations’, eLife, p. Forthcoming, Jun. 2022, doi: 10.1101/2022.06.16.22276024.

[17] C. Viboud et al., ‘The RAPIDD ebola forecasting challenge: Synthesis and lessons learnt’, Epidemics, vol. 22, pp. 13–21, Mar. 2018, doi: 10.1016/j.epidem.2017.08.002.

[18] J. Bracher et al., ‘A pre-registered short-term forecasting study of COVID-19 in Germany and Poland during the second wave’, Nat. Commun., vol. 12, no. 1, p. 5173, Aug. 2021, doi: 10.1038/s41467-021-25207-0.

[19] E. Y. Cramer et al., ‘Evaluation of individual and ensemble probabilistic forecasts of COVID-19 mortality in the United States’, Proc. Natl. Acad. Sci., vol. 119, no. 15, p. e2113561119, Apr. 2022, doi: 10.1073/pnas.2113561119.

[20] J. W. Taylor and K. S. Taylor, ‘Combining Probabilistic Forecasts of COVID-19 Mortality in the United States’, Eur. J. Oper. Res., Jun. 2021, doi: 10.1016/j.ejor.2021.06.044.

[21] E. Howerton et al., ‘Context-dependent representation of within- and between-model uncertainty: aggregating probabilistic predictions in infectious disease epidemiology’, J. R. Soc. Interface, vol. 20, no. 198, p. 20220659, Jan. 2023, doi: 10.1098/rsif.2022.0659.

[22] C. Genest, ‘Vincentization Revisited’, Ann. Stat., vol. 20, no. 2, pp. 1137–1142, 1992.

[23] J. L. Juul, K. Græsbøll, L. E. Christiansen, and S. Lehmann, ‘Fixed-time descriptive statistics underestimate extremes of epidemic curve ensembles’, Nat. Phys., vol. 17, no. 1, Art. no. 1, Jan. 2021, doi: 10.1038/s41567-020-01121-y.

[24] E. Dong, H. Du, and L. Gardner, ‘An interactive web-based dashboard to track COVID-19 in real time’, Lancet Infect. Dis., vol. 20, no. 5, pp. 533–534, May 2020, doi:10.1016/S1473-3099(20)30120-1.

[25] European COVID-19 Scenario Hub, ‘Round 2’. [Online]. Available: https://covid19scenariohub.eu/report2.html

[26] K. Sherratt and S. Funk, ‘covid19-forecast-hub-europe/aggregation-info-loss’. [Online]. Available: https://github.com/covid19-forecast-hub-europe/aggregation-info-loss/releases/

[27] Pedro J. Aphalo, ‘ggpmisc: Miscellaneous Extensions to “ggplot2”‘. 2023. [Online]. Available: https://docs.r4photobiology.info/ggpmisc/

[28] F. E. Harrell and C. E. Davis, ‘A new distribution-free quantile estimator’, Biometrika, vol. 69, no. 3, pp. 635–640, Dec. 1982, doi: 10.1093/biomet/69.3.635.

[29] A. Lenhard, W. Lenhard, and S. Gary, ‘cNORM - Generating Continuous Test Norms’. 2018. doi: 10.13140/RG.2.2.25821.26082.

[30] N. I. Bosse, S. Abbott, A. Cori, E. van Leeuwen, J. Bracher, and S. Funk, ‘Scoring epidemiological forecasts on transformed scales’, PLOS Comput. Biol., vol. 19, no. 8, p. e1011393, Aug. 2023, doi: 10.1371/journal.pcbi.1011393.

[31] K. Sherratt et al., ‘Improving modelling for epidemic responses: reflections from members of the UK infectious disease modelling community on their experiences during the COVID-19 pandemic [version 1; peer review: awaiting peer review]’, Wellcome Open Res., vol. 9, no. 12, 2024, doi: 10.12688/wellcomeopenres.19601.1.

[32] A. Srivastava, S. Singh, and F. Lee, ‘Shape-based Evaluation of Epidemic Forecasts’. arXiv, Nov. 11, 2022. doi: 10.48550/arXiv.2209.04035.

