## Supplement for "Characterising information gains and losses when collecting multiple epidemic model outputs"

### Supplementary Information

#### SI Figure 1

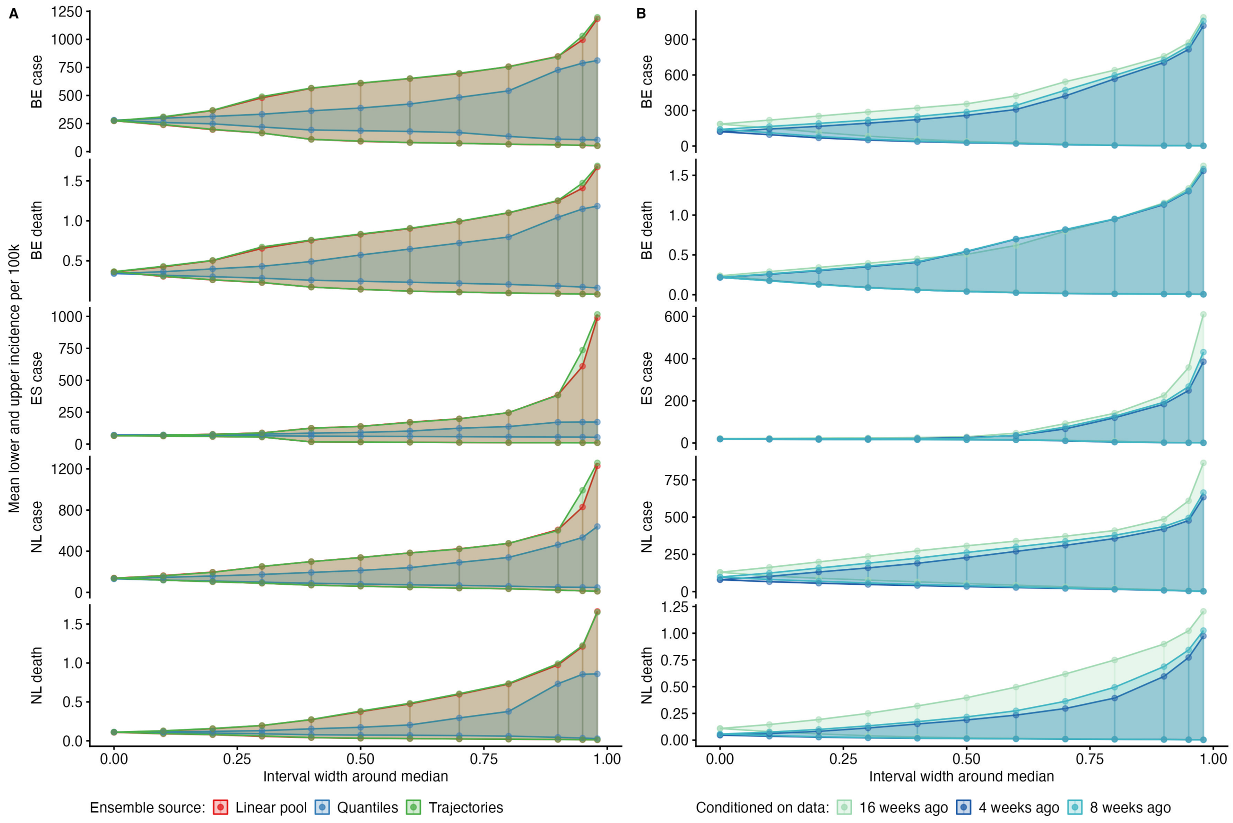


SI Figure 1. Mean central prediction intervals at increasing distances from the median. The 52-week mean of incidence per 100,000 population across all time points and scenarios, showing mean central prediction intervals at increasing distances from the median (interval width), by aggregation method (A) or weighting (B). The median estimate for each ensemble has 0 interval width (x-axis), with uncertainty increasing until an interval width at 0.98 represents the 1%-99% credibility interval around the median.

#### SI Figure 2

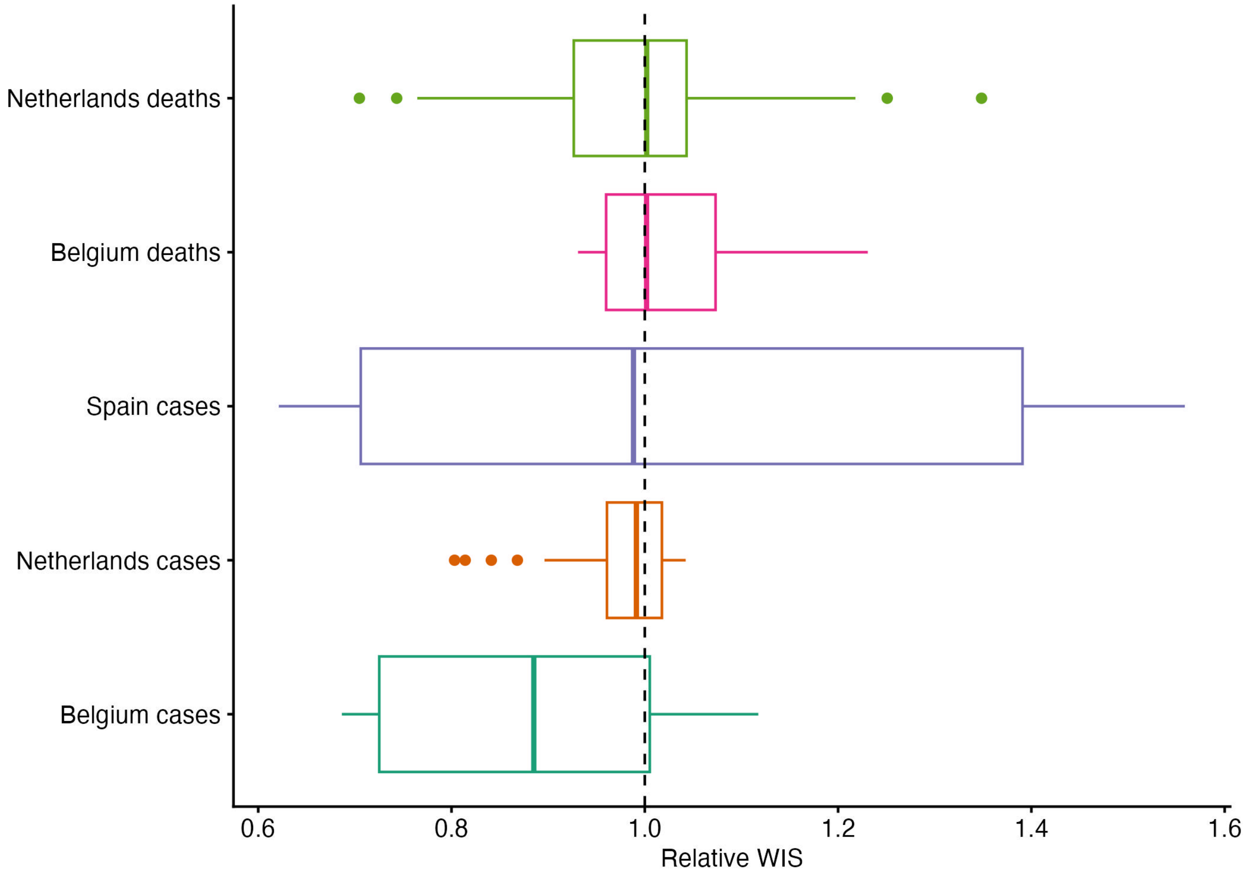


SI Figure 2. Distribution of forecast performance scores (relative WIS), of forecasts from model trajectories weighted using 0 through 31 weeks' available data. Performance is compared to an unweighted ensemble (reference line at 1).

#### SI Table 1

| Team | Methods |
|---|---|
| **ECDC** | |
| ECDC-CM_ONE | Discrete-time, deterministic, mean-field SEIR-type compartmental model on metapopulation level. Population divided by age, vaccination status, and previous recovery; incl. seasonality, BA2 & behavior. |
| **Dutch National Institute of Public Health and the Environment (RIVM)** | |
| RIVM-vacamole | Deterministic, age-structured SEIR model, accounting for differences in susceptibility/infectiousness by age, seasonality, contact patterns, modes of vaccine protection, and waning immunity. |
| **SIMID** | |
| SIMID-SCM | Stochastic age-structured discrete time extended compartmental model |
| **Universidad Carlos III de Madrid** | |

| Team | Methods |
| --- | --- |
| UC3M-EpiGraph | Agent-based parallel simulator that models individual interactions extracted from social networks and demographical data. |
| **University of Southern California** | |
| USC-SIkJalpha | Uses SIKJalpha which models temporally varying infection, death, and hospitalization rates. Learning is performed by reducing the problem to multiple simple linear regression problems. |

SI Table 1. Teams that contributed models to Round 2 of the European Scenario Hub, with self-described methods and links to further information. See also:

- Full model metadata, at: https://github.com/covid19-forecast-hub-europe/covid19-scenario-hub-europe/tree/main/model-metadata
- Information about each model's assumptions for Round 2, at: https://github.com/covid19-forecast-hub-europe/covid19-scenario-hub-europe/tree/main/model-abstracts/2022-07-24

### Round 2 report

The following pages are the original website reporting for the European Scenario Hub Round 2 as of July 2022.

The report is currently (January 2023) available at: https://covid19scenariohub.eu/report2.html

Code to generate this report is available at: https://github.com/european-modelling-hubs/covid19-scenario-hub-europe-website/blob/main/report2.Rmd

### Round 2

#### Scenarios

We asked teams of researchers across Europe to use quantitative models to project COVID-19 outcomes for 32 European countries over the next year. In order to explore different sets of assumptions about drivers of the pandemic, we asked teams to vary four sets of parameters. We can describe this in a 2x2 scenario specification:

<table>
<tr><td></td><td>Age 60+ booster campaign<ul><li>2nd* booster recommended for 60+</li><li>Uptake starts 15th September, and reaches 50% coverage by 15th December</li></ul></td><td>Age 18+ booster campaign<ul><li>2nd* booster recommended for general population, ages 18+</li><li>Uptake starts 15th September, and reaches 50% coverage by 15th December</li></ul></td></tr>
<tr><td>Optimistic vaccine effectiveness<ul><li>Increased booster vaccine effectiveness to that seen against Delta variant</li></ul></td><td>Scenario A</td><td>Scenario B</td></tr>
<tr><td>Pessimistic vaccine effectiveness<ul><li>Reduced booster vaccine effectiveness against infection from BA.4/BA.5/BA.2.75 variants</li></ul></td><td>Scenario C</td><td>Scenario D</td></tr>
</table>

See also the full scenario details (https://github.com/covid19-forecast-hub-europe/covid19-scenario-hub-europe/wiki/Round-2) for more detail on the common set of assumptions teams used to create their models.

In Round 2, we asked modellers to start their projections from the 2022-07-24. Data after this date were not included, and as a result, model projections are unlikely to fully account for later information on the changing variants or behavioural patterns.

In this report we only show results from countries with at least 3 models.

#### Current situation

We consider vaccination rates in countries for which multiple teams of modellers contributed projections.

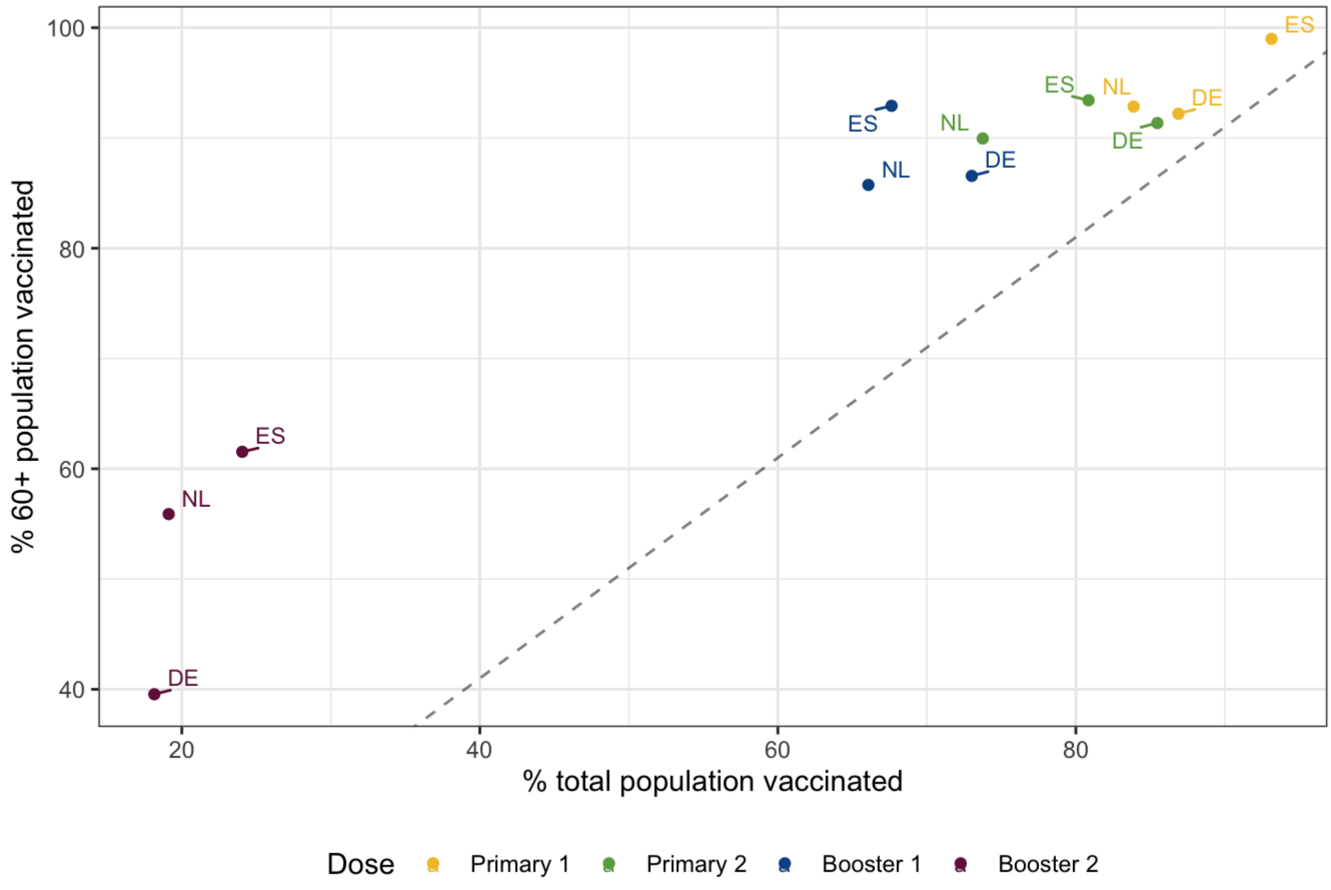

Vaccination % among 60+ population, relative to total population

### Participating teams

6 models contributed scenario projections to Round 2.

#### Models

Participating teams by number of countries and horizon

| Team | Countries | Weeks |
|---|---|---|
| USC-SIkJalpha | 31 | 52 |
| ECDC-CM_ONE | 28 | 53 |
| MODUS_Covid-Episim | 1 | 53 |
| RIVM-vacamole | 1 | 53 |
| SIMID-SCM | 1 | 52 |
| UC3M-EpiGraph | 1 | 41 |

#### Countries

Number of independent model projections for each target variable and location

| Code | Country | Infection | Case | Hosp | Icu | Death |
|---|---|---|---|---|---|---|
| BE | Belgium | 1 | 3 | 2 | 1 | 3 |
| DE | Germany | 1 | 2 | 2 | 0 | 1 |
| ES | Spain | 1 | 3 | 2 | 0 | 2 |

| Code | Country | Infection | Case | Hosp | Icu | Death |
|---|---|---|---|---|---|---|
| NL | Netherlands | 1 | 3 | 2 | 1 | 3 |

### Cumulative outcomes

For each model and scenario, we compare the total number of outcomes over the entire projection period as a % of the total country population. We compared the cumulative number of projected outcomes to the cumulative total over one year before projections started (July 2021 to July 2022).

#### Death

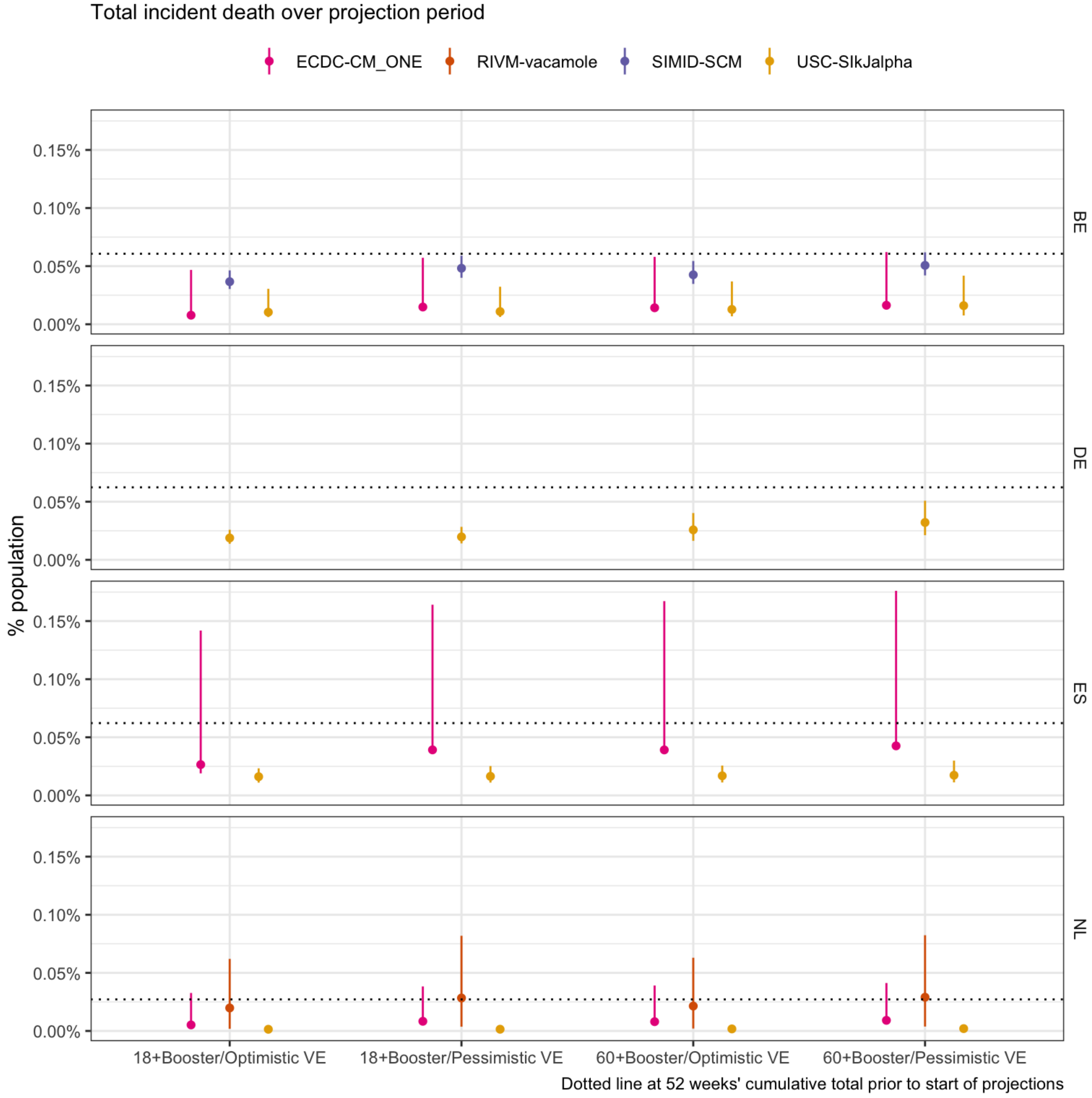


Scenarios: Autumn second booster campaign among population aged ‘18+’ or ‘60+’; Vaccine effectiveness is ‘optimistic’(effectiveness as of a booster vaccine against Delta) or ‘pessimistic’ (as against BA.4/BA.5/BA.2.75)

### Case

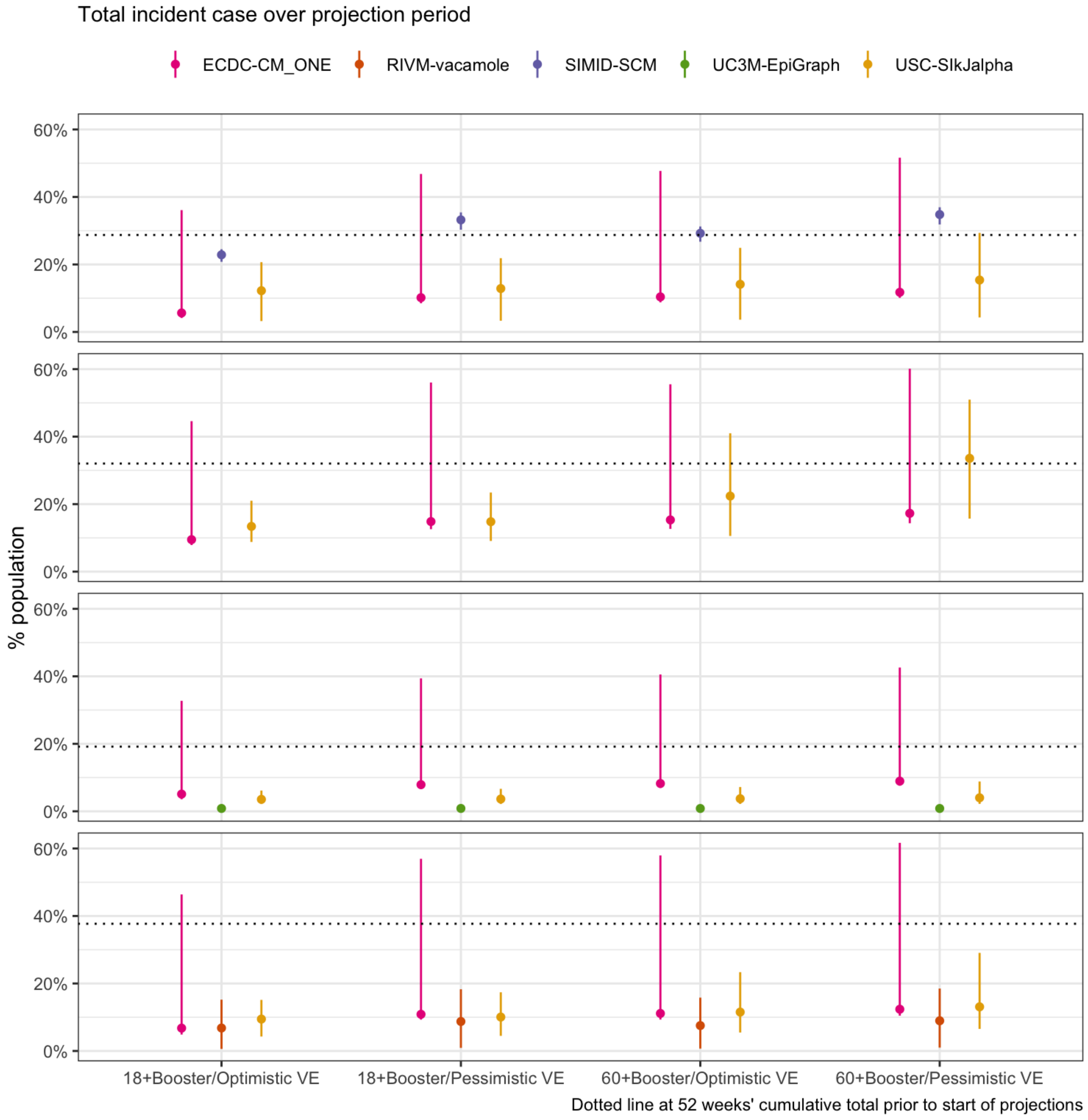


Scenarios: Autumn second booster campaign among population aged ‘18+’ or ‘60+’; Vaccine effectiveness is ‘optimistic’(effectiveness as of a booster vaccine against Delta) or ‘pessimistic’ (as against BA.4/BA.5/BA.2.75)

### Infection

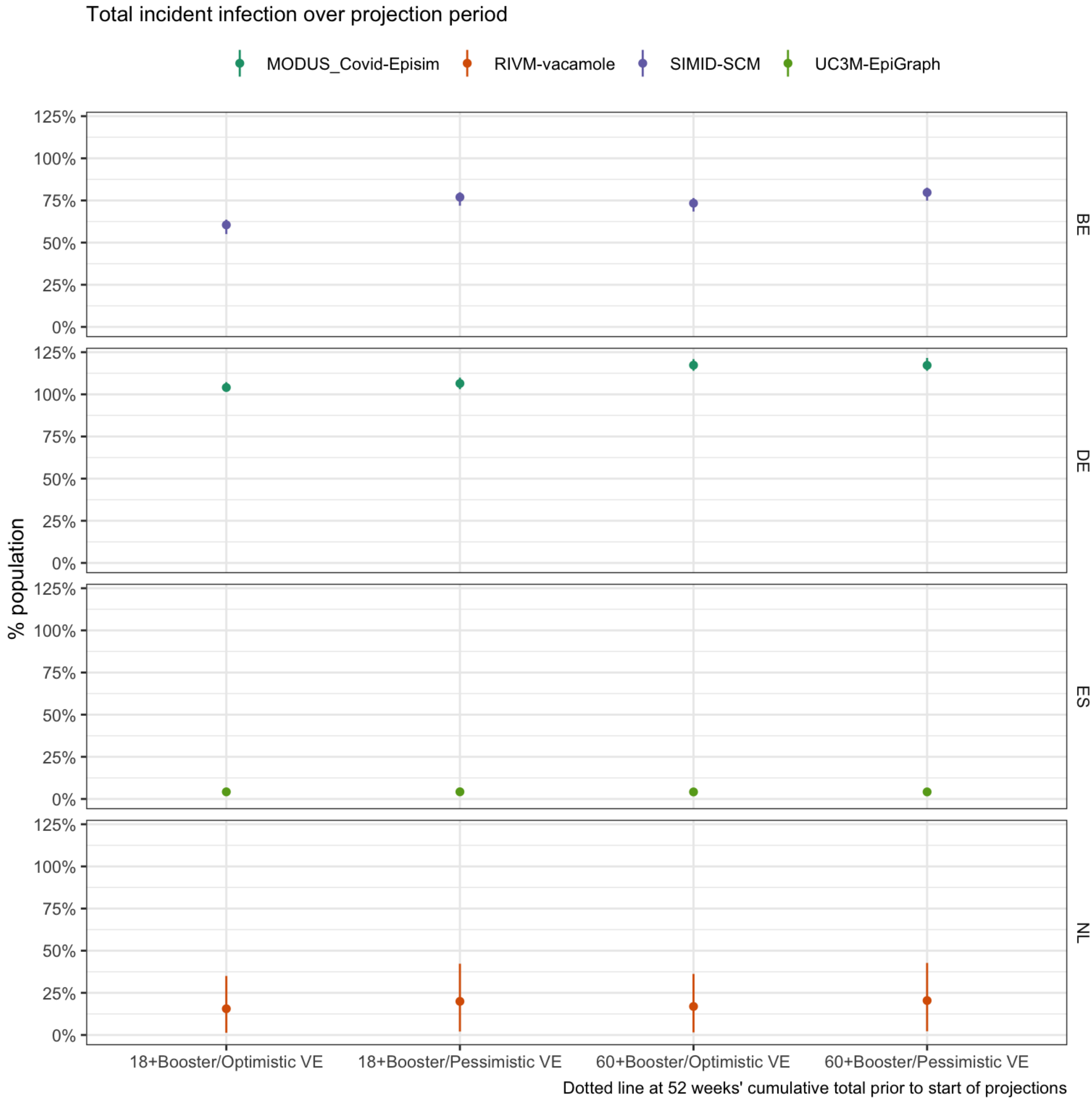


Scenarios: Autumn second booster campaign among population aged ‘18+’ or ‘60+’; Vaccine effectiveness is ‘optimistic’(effectiveness as of a booster vaccine against Delta) or ‘pessimistic’ (as against BA.4/BA.5/BA.2.75)

### Hosp

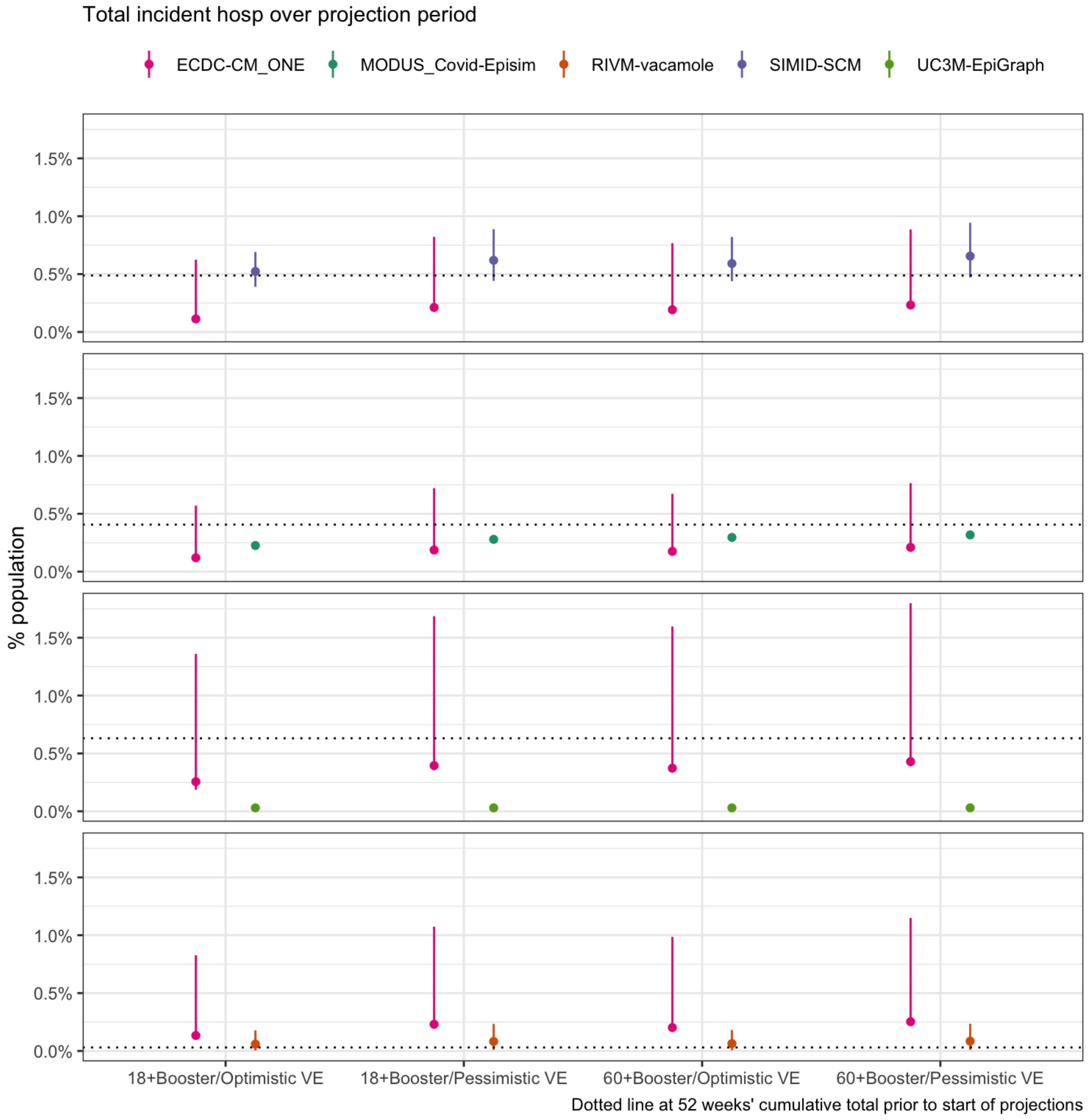


Scenarios: Autumn second booster campaign among population aged ‘18+’ or ‘60+’; Vaccine effectiveness is ‘optimistic’(effectiveness as of a booster vaccine against Delta) or ‘pessimistic’ (as against BA.4/BA.5/BA.2.75)

### Icu

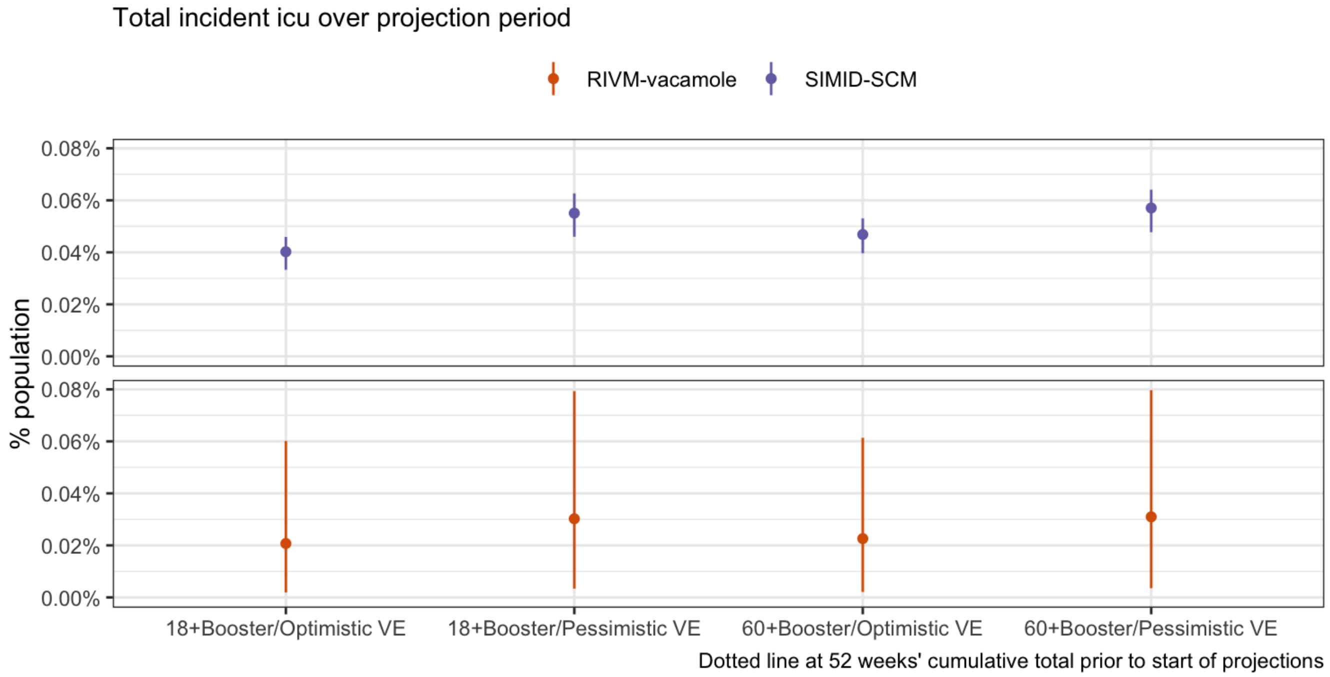


Scenarios: Autumn second booster campaign among population aged '18+' or '60+'; Vaccine effectiveness is 'optimistic'(effectiveness as of a booster vaccine against Delta) or 'pessimistic' (as against BA.4/BA.5/BA.2.75)

### Incident outcomes

We explored the incidence of COVID-19 per 100,000 over the projection period and in terms of projected peaks in incidence. We summarised peaks both over the entire projection period, and over only the autumn-winter period (October through March); we considered (A) the timing and maximum weekly incidence of each peak, and (B) the total number of peaks.

### Trajectories

#### Death

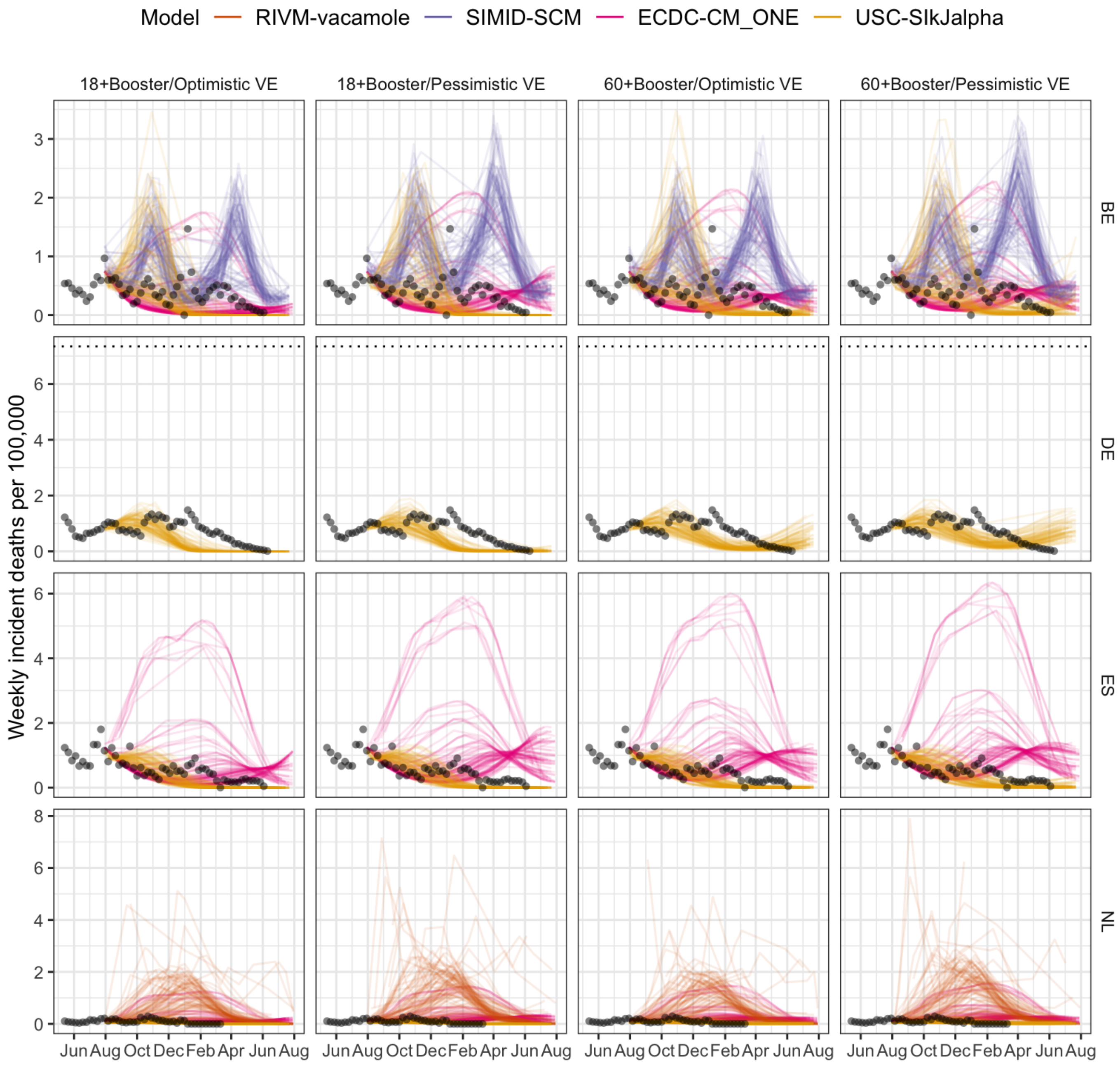


Scenarios: Autumn second booster campaign among population aged '18+' or '60+';
Vaccine effectiveness is 'optimistic'(effectiveness as of a booster vaccine against Delta) or 'pessimistic' (as against BA.4/BA.5/BA.2.75)

### Case

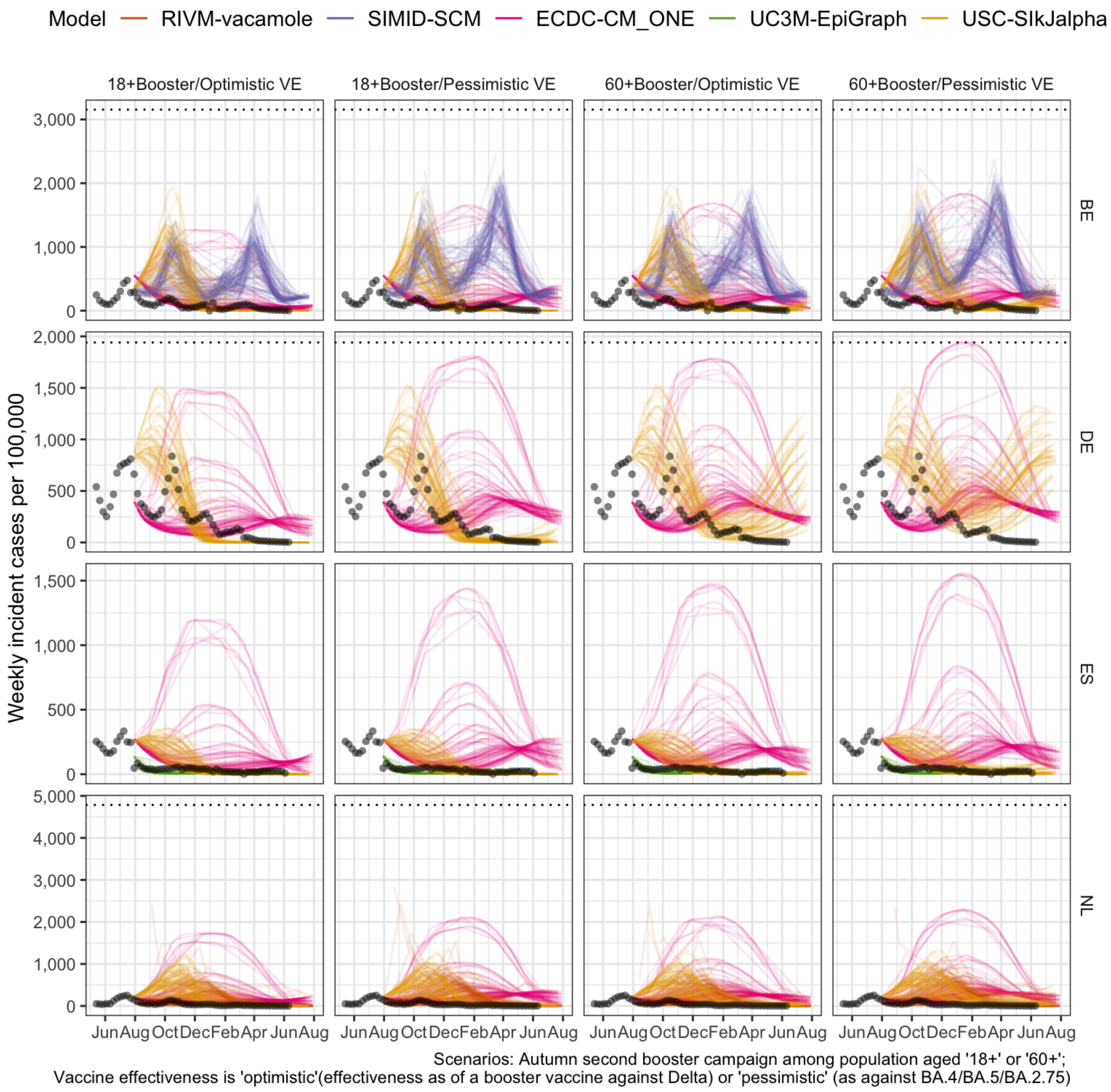

Model
RIVM-vacamole
SIMID-SCM
ECDC-CM_ONE
UC3M-EpiGraph
USC-SIkJalpha
18+Booster/Optimistic VE
18+Booster/Pessimistic VE
60+Booster/Optimistic VE
60+Booster/Pessimistic VE
BE
DE
ES
NL
Weekly incident cases per 100,000
Jun Aug Oct Dec Feb Apr Jun Aug
Scenarios: Autumn second booster campaign among population aged '18+' or '60+';
Vaccine effectiveness is 'optimistic'(effectiveness as of a booster vaccine against Delta) or 'pessimistic' (as against BA.4/BA.5/BA.2.75)

### Infection

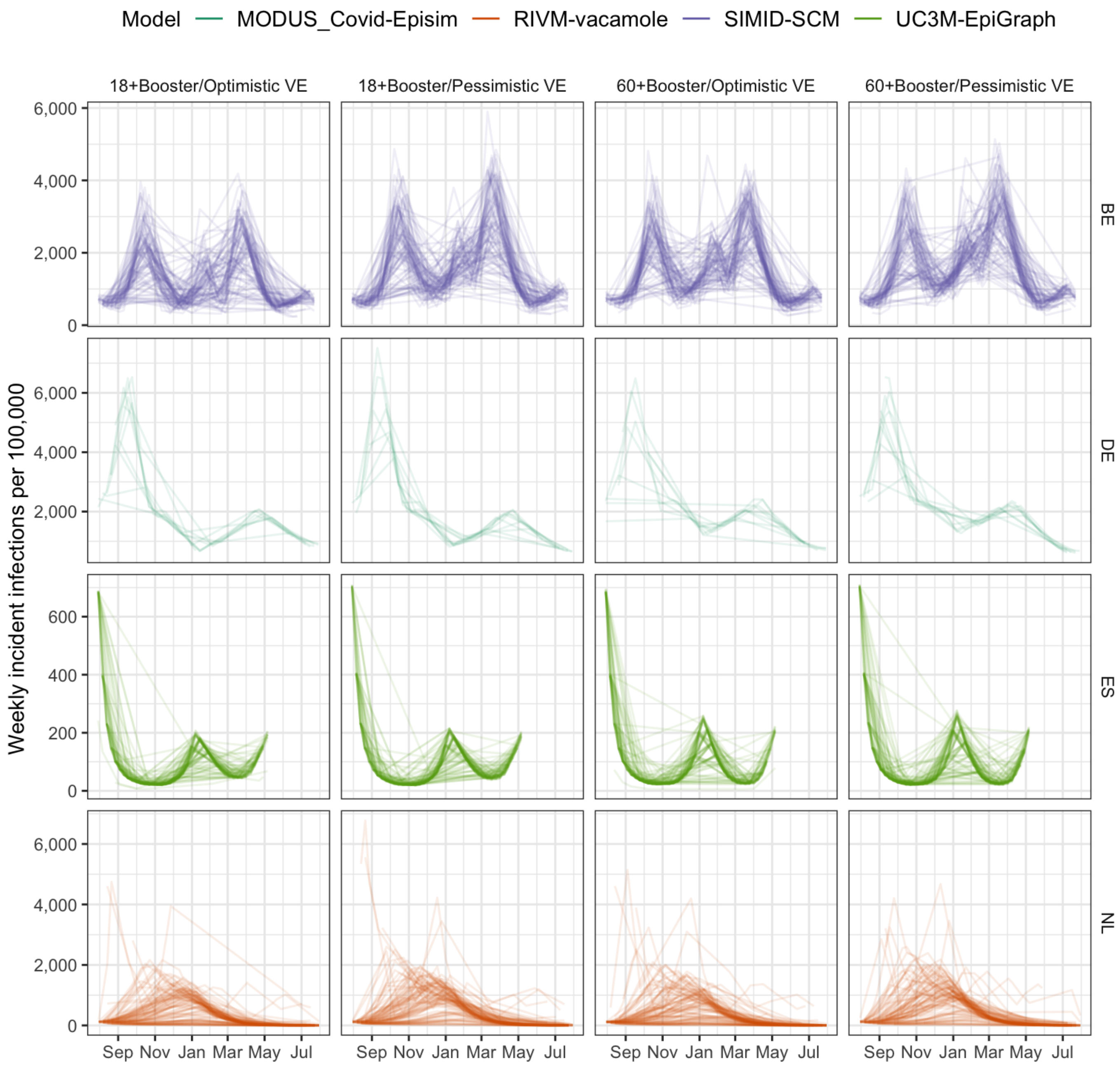


Scenarios: Autumn second booster campaign among population aged '18+' or '60+';
Vaccine effectiveness is 'optimistic'(effectiveness as of a booster vaccine against Delta) or 'pessimistic' (as against BA.4/BA.5/BA.2.75)

### Hosp

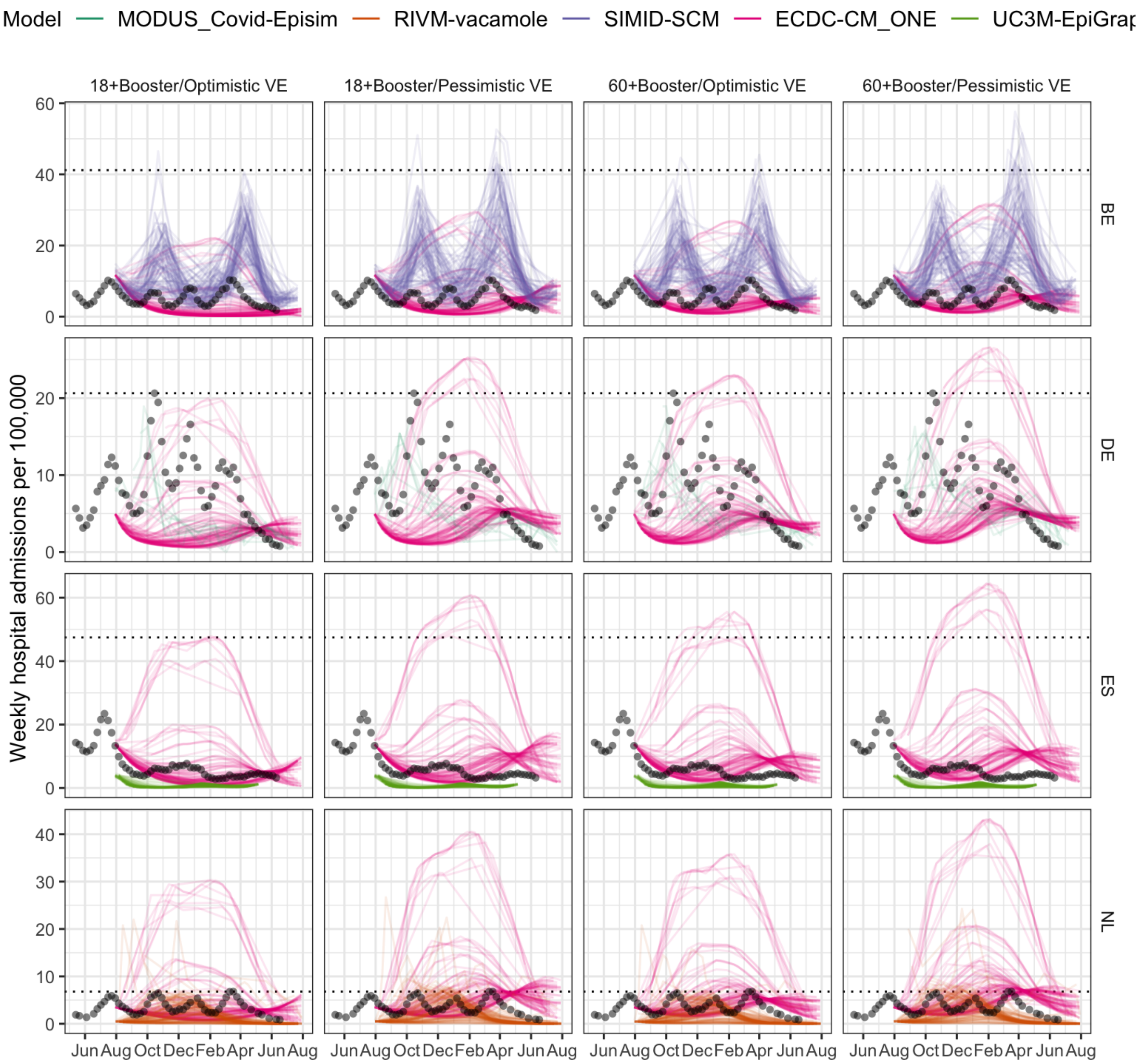


Scenarios: Autumn second booster campaign among population aged '18+' or '60+';
Vaccine effectiveness is 'optimistic'(effectiveness as of a booster vaccine against Delta) or 'pessimistic' (as against BA.4/BA.5/BA.2.75)

##### Icu

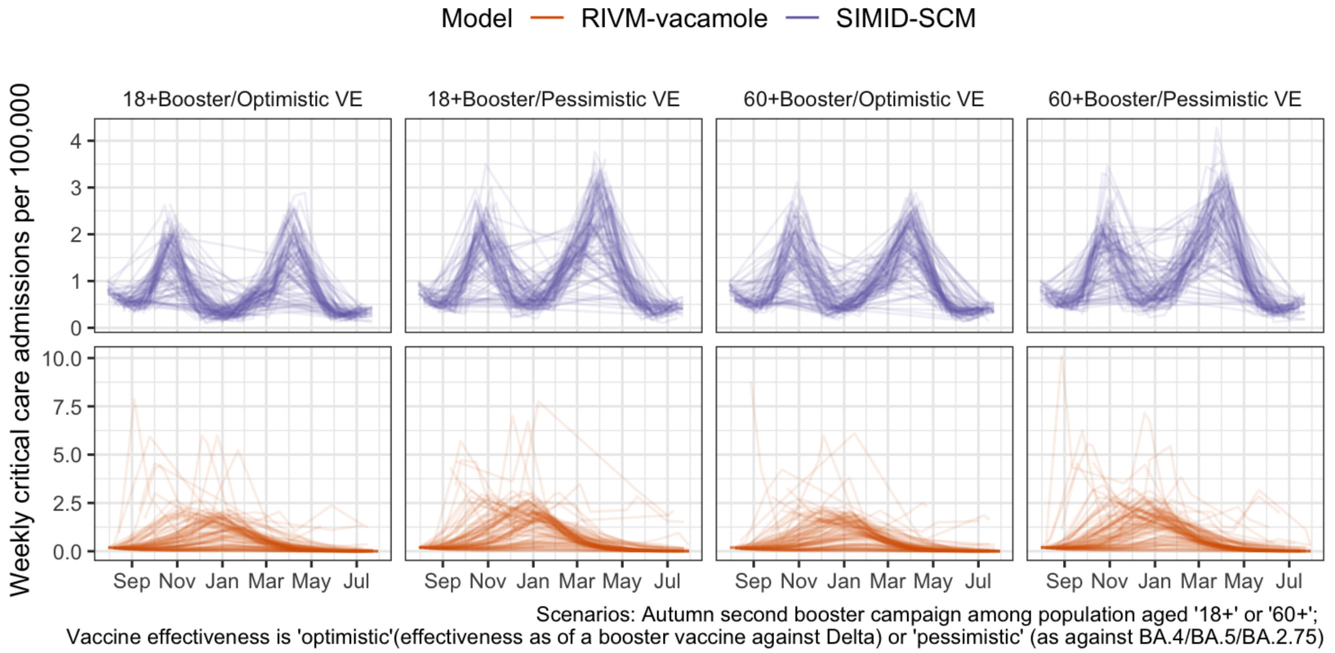


### Peaks

#### Autumn-winter

*Projections over October 2022 through March 2023*

##### Death

A. Size and timing of peaks. Boxplots show summary of the likely value at peak incidence (median and interquartile range); points show timing and size of peaks from independent sample simulations

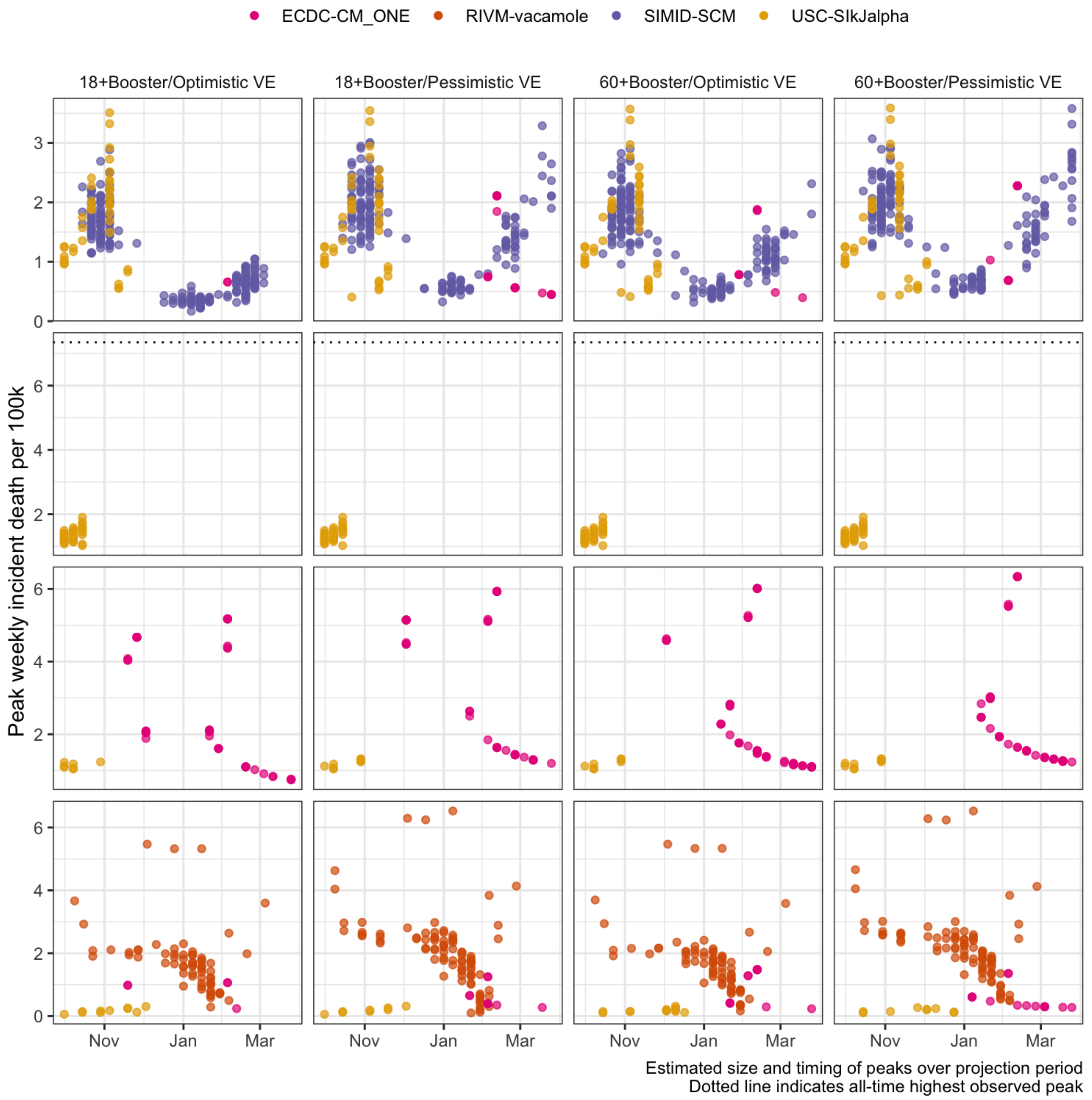


B. Projected number of peaks (median with 5-95% probability)

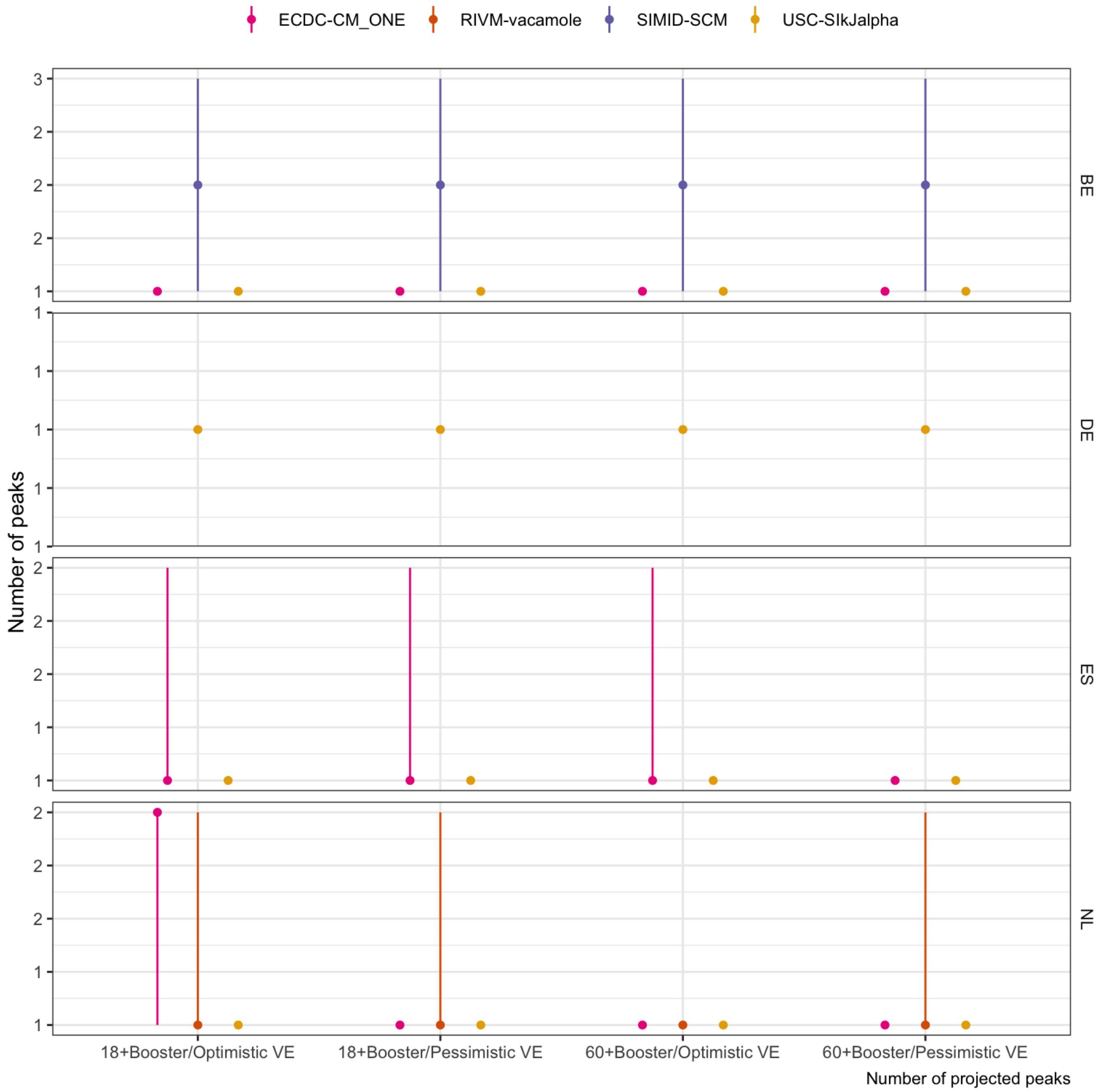


Scenarios: Autumn second booster campaign among population aged ‘18+’ or ‘60+’; Vaccine effectiveness is ‘optimistic’(effectiveness as of a booster vaccine against Delta) or ‘pessimistic’ (as against BA.4/BA.5/BA.2.75)

#### Case

A. Size and timing of peaks. Boxplots show summary of the likely value at peak incidence (median and interquartile range); points show timing and size of peaks from independent sample simulations

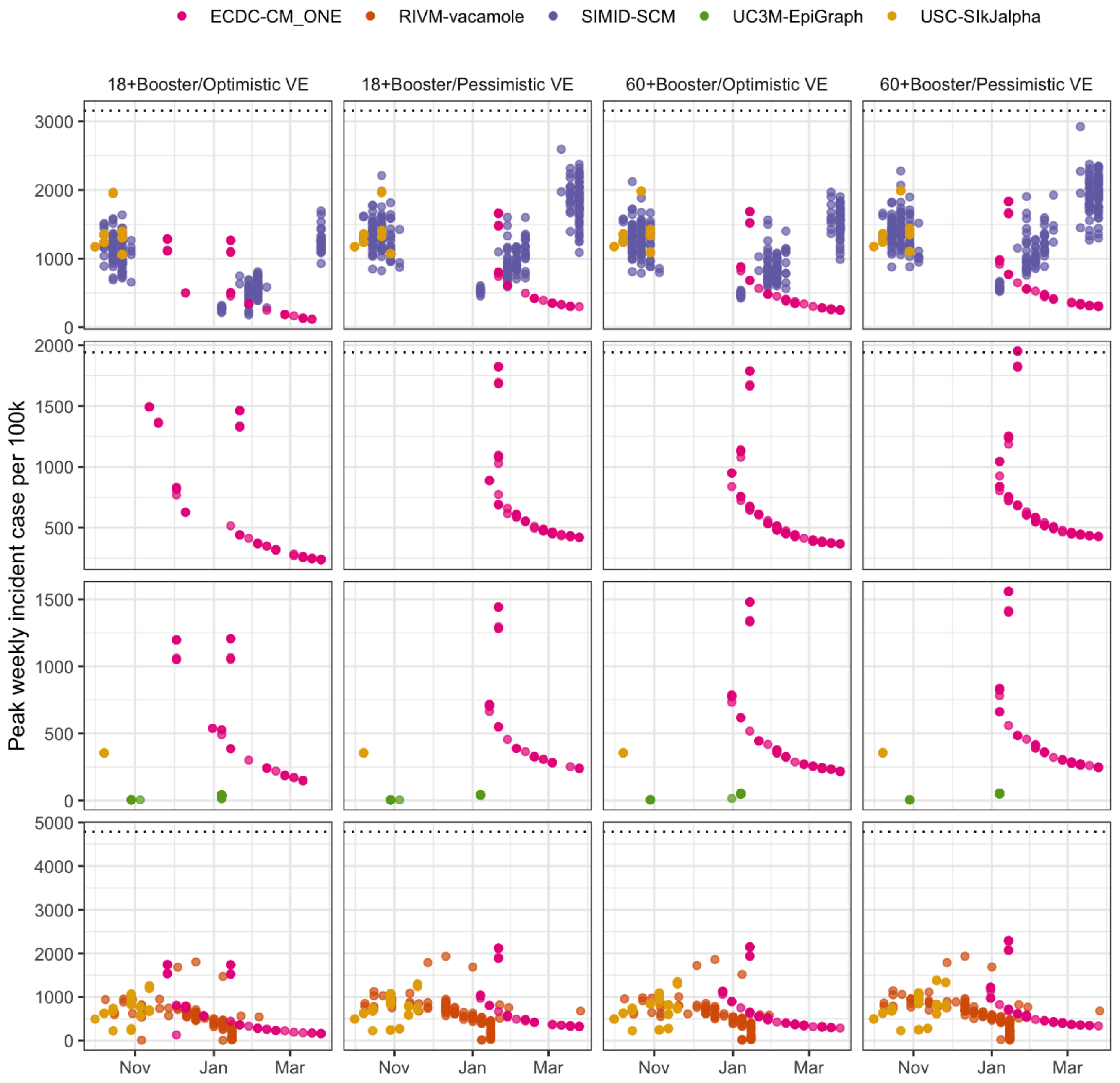


Estimated size and timing of peaks over projection period
Dotted line indicates all-time highest observed peak

B. Projected number of peaks (median with 5-95% probability)

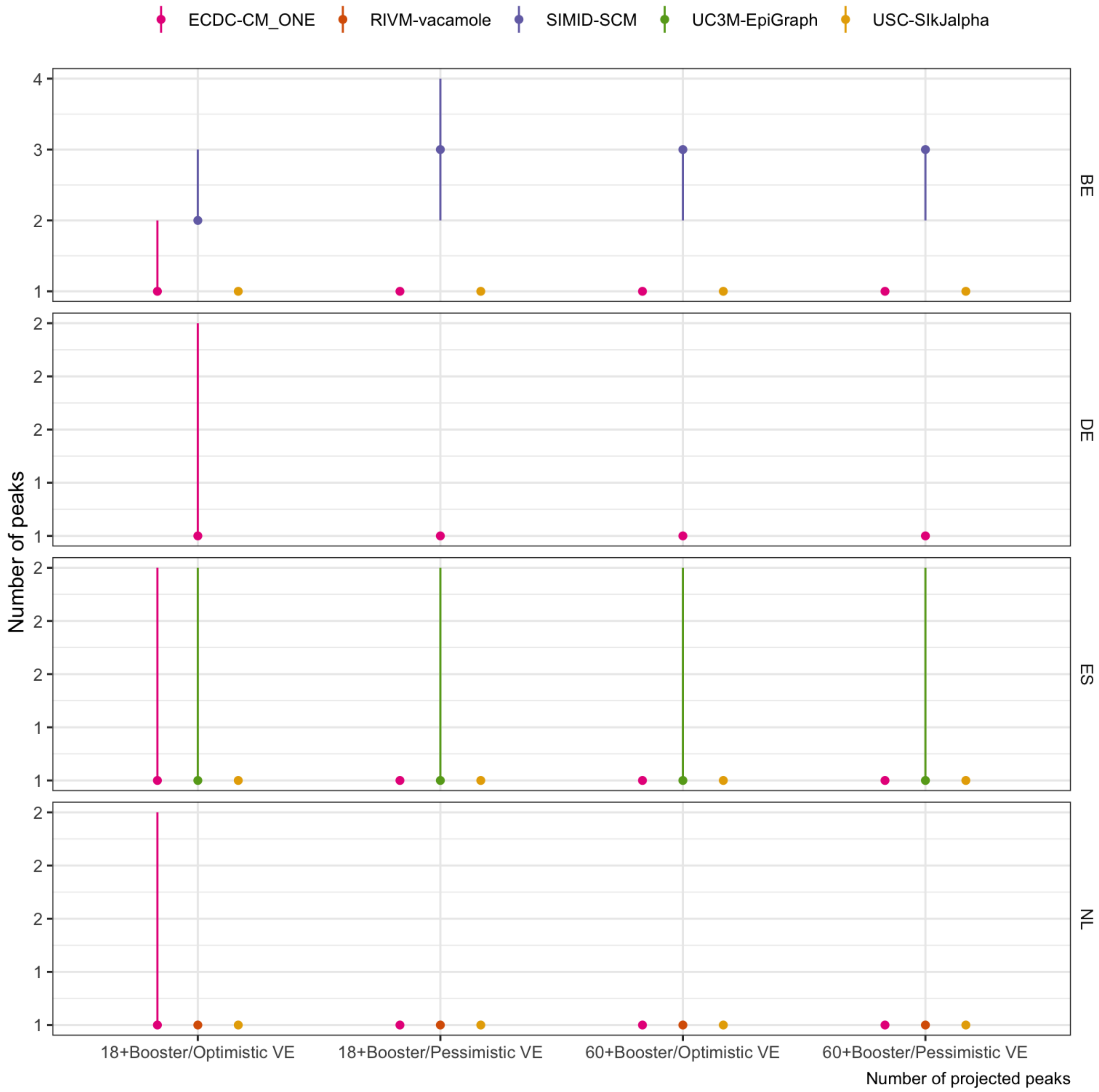


Scenarios: Autumn second booster campaign among population aged ‘18+’ or ‘60+’; Vaccine effectiveness is ‘optimistic’(effectiveness as of a booster vaccine against Delta) or ‘pessimistic’ (as against BA.4/BA.5/BA.2.75)

#### Infection

A. Size and timing of peaks. Boxplots show summary of the likely value at peak incidence (median and interquartile range); points show timing and size of peaks from independent sample simulations

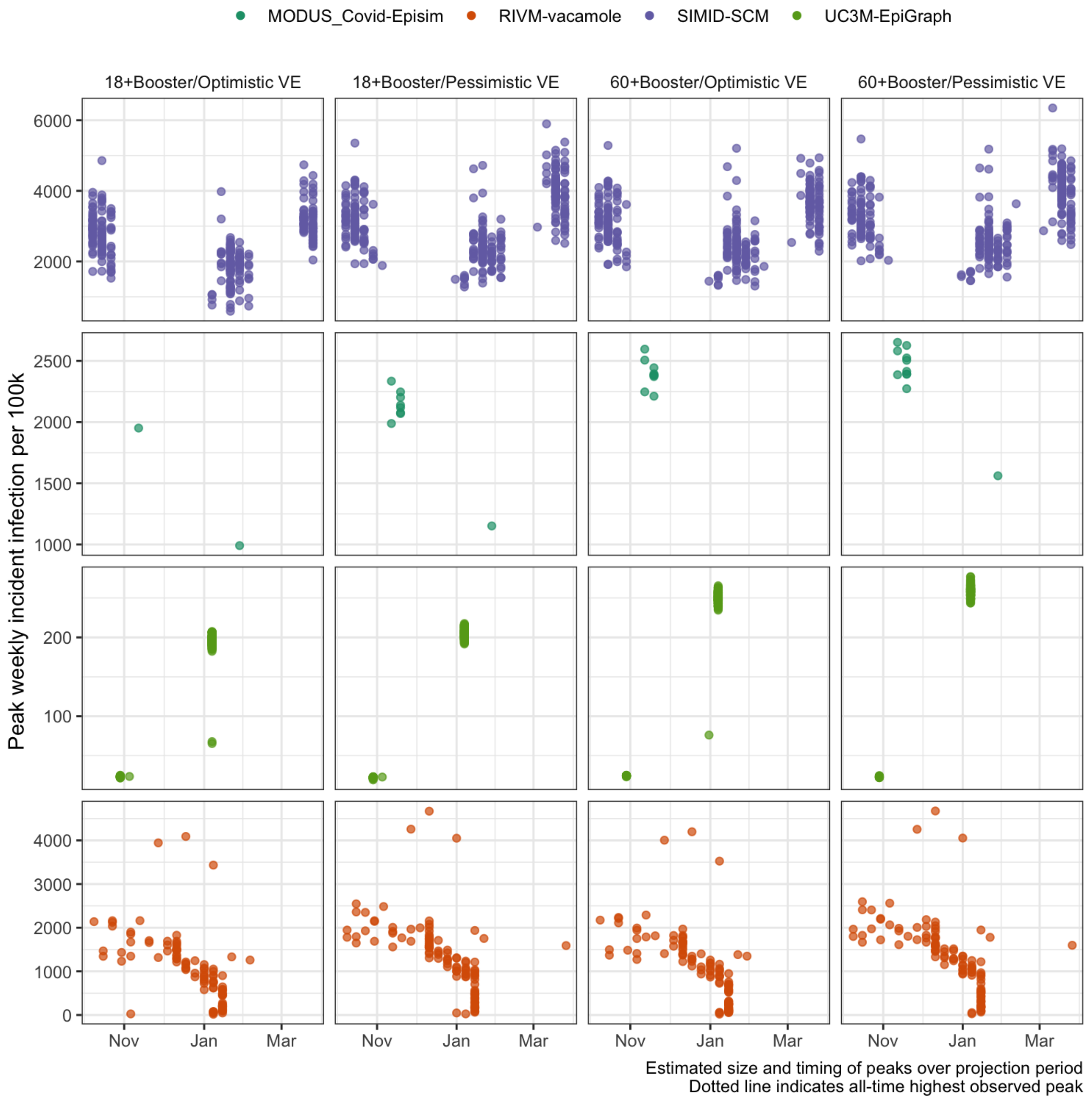


B. Projected number of peaks (median with 5-95% probability)

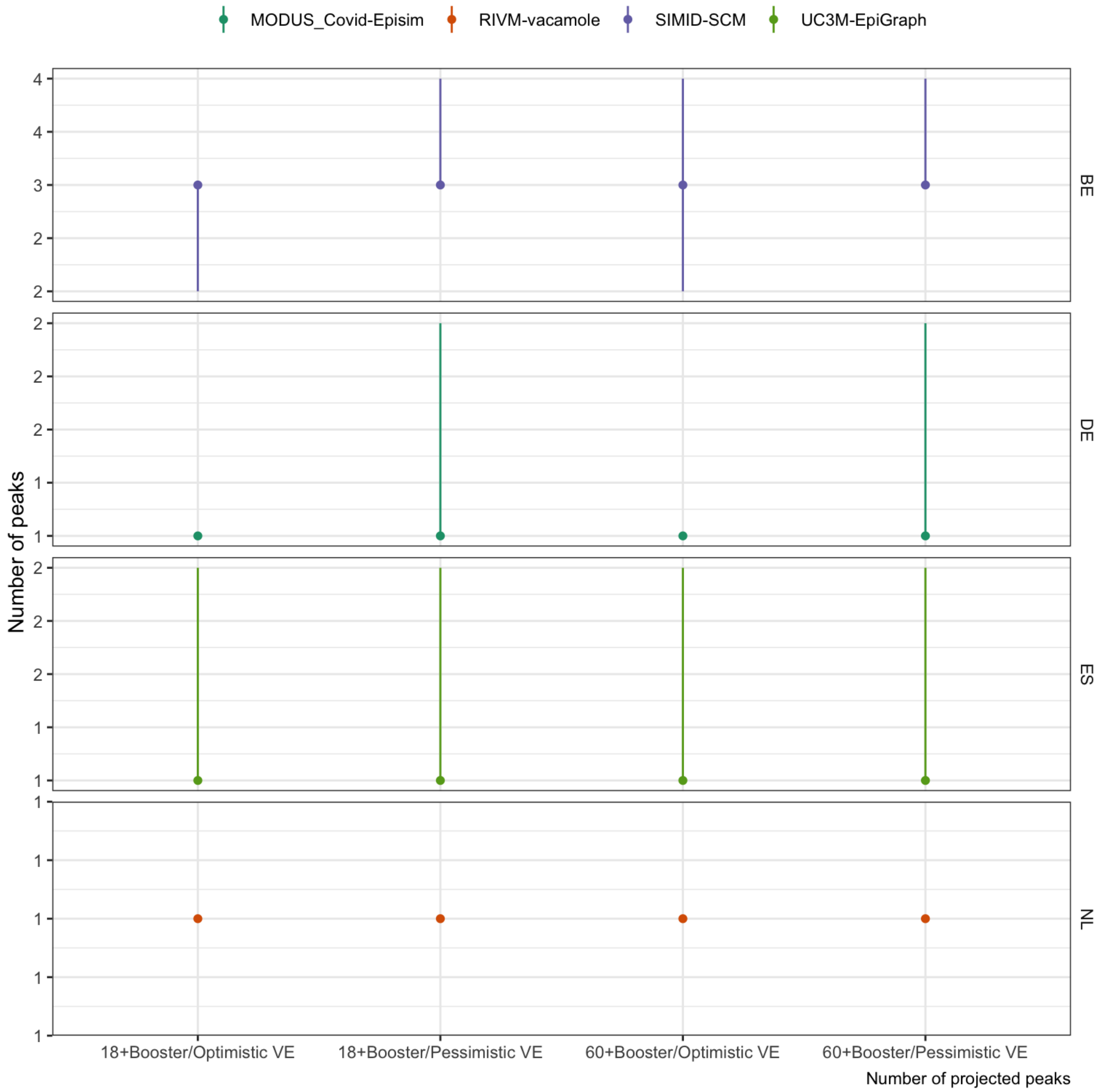


Scenarios: Autumn second booster campaign among population aged '18+' or '60+'; Vaccine effectiveness is 'optimistic'(effectiveness as of a booster vaccine against Delta) or 'pessimistic' (as against BA.4/BA.5/BA.2.75)

#### Hosp

A. Size and timing of peaks. Boxplots show summary of the likely value at peak incidence (median and interquartile range); points show timing and size of peaks from independent sample simulations

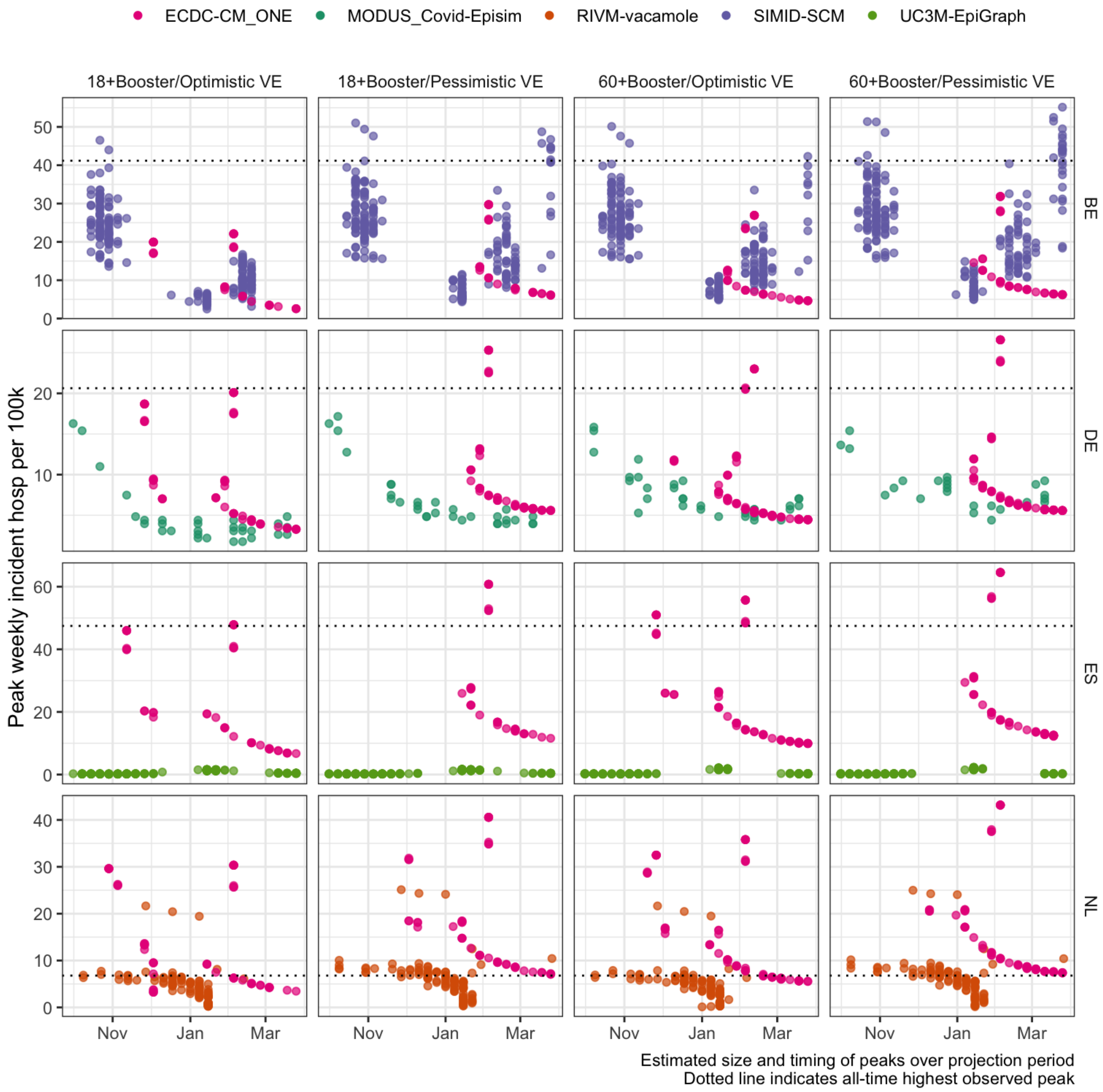


B. Projected number of peaks (median with 5-95% probability)

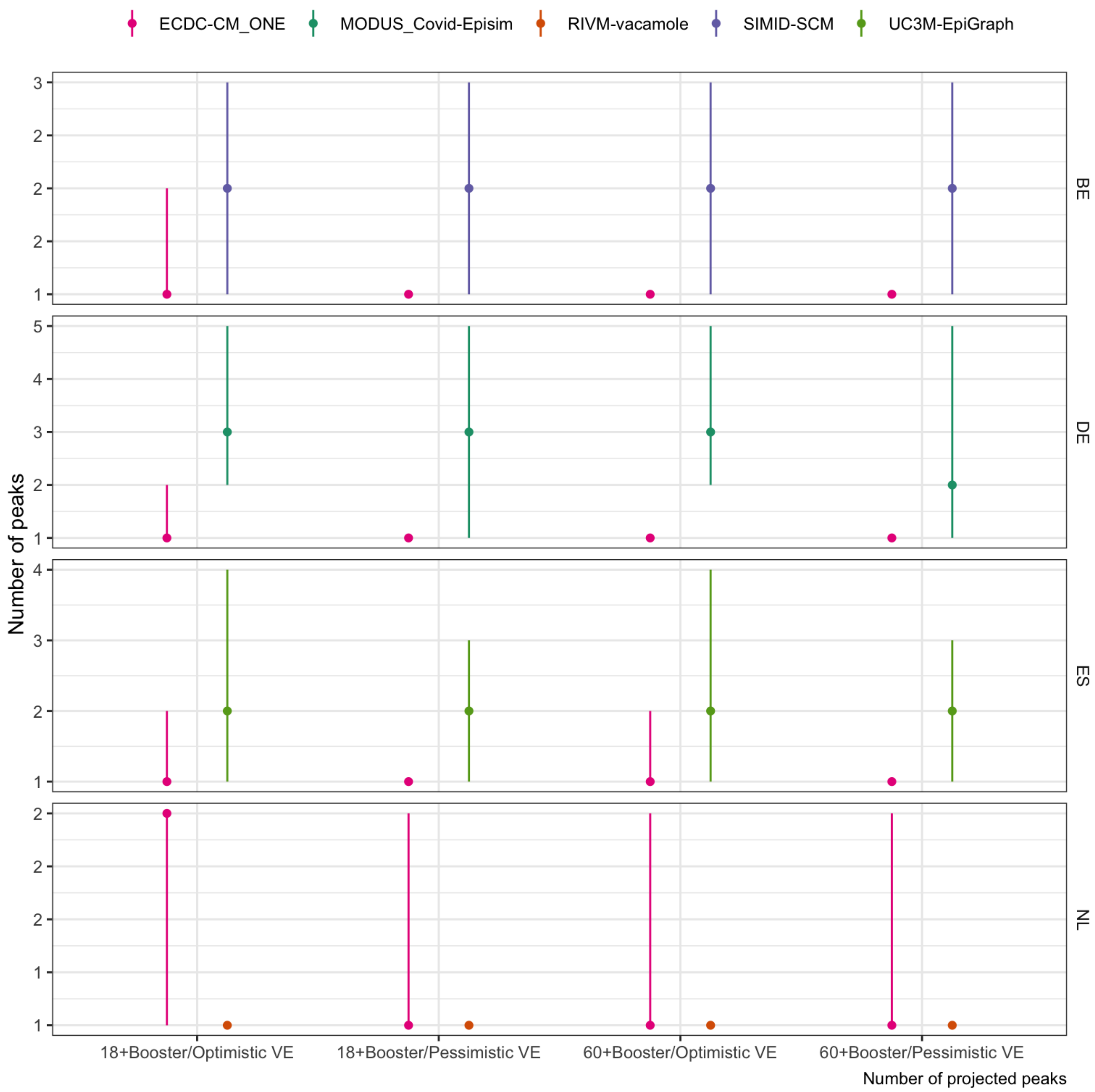


Scenarios: Autumn second booster campaign among population aged ‘18+’ or ‘60+’; Vaccine effectiveness is ‘optimistic’(effectiveness as of a booster vaccine against Delta) or ‘pessimistic’ (as against BA.4/BA.5/BA.2.75)

Icu

A. Size and timing of peaks. Boxplots show summary of the likely value at peak incidence (median and interquartile range); points show timing and size of peaks from independent sample simulations

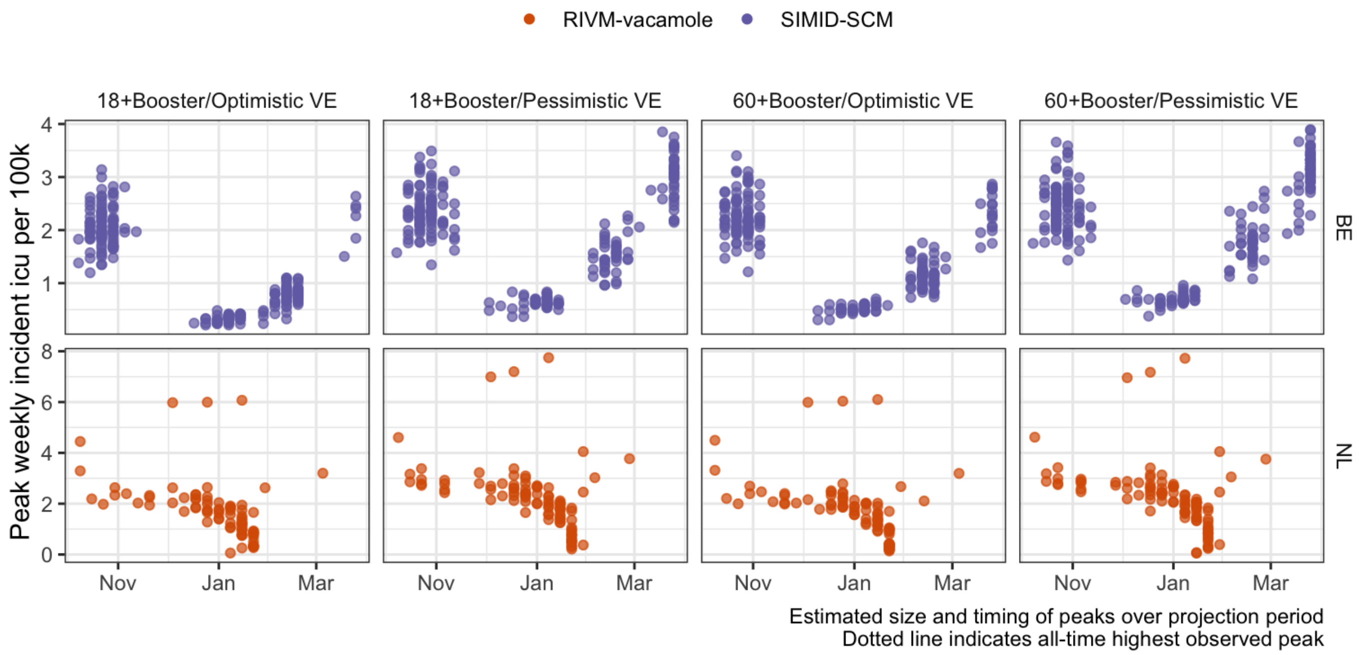


B. Projected number of peaks (median with 5-95% probability)

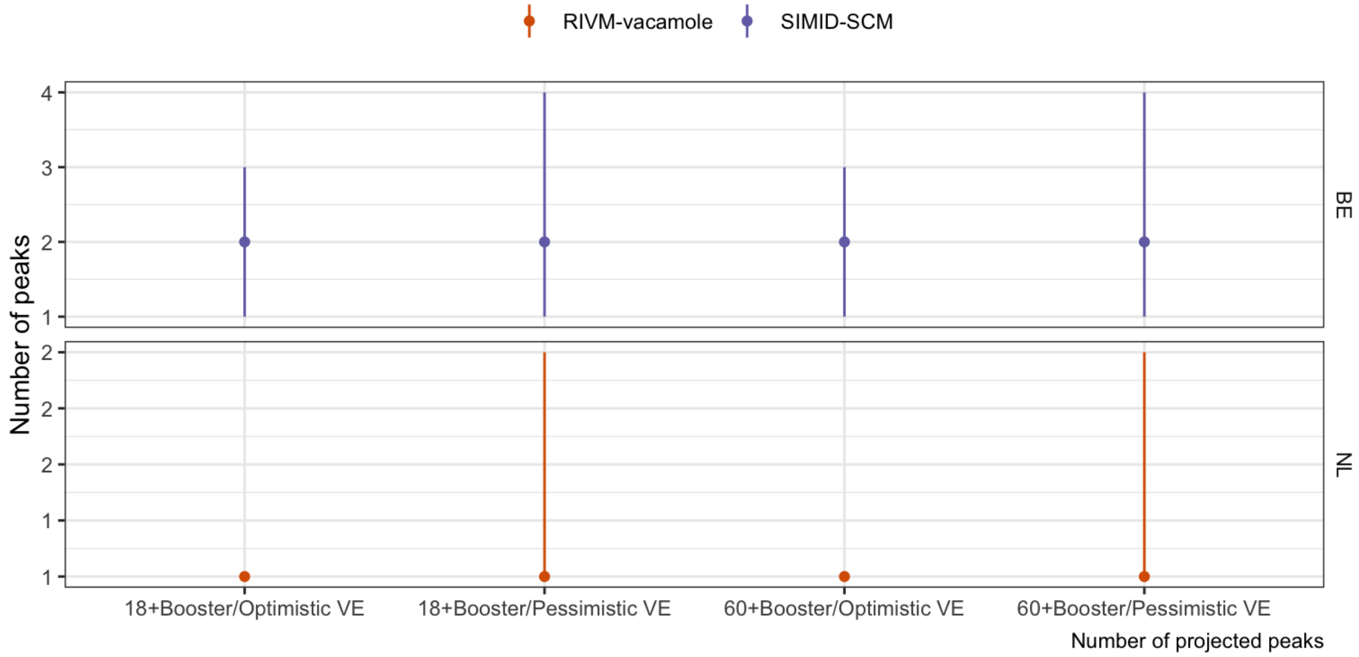


Scenarios: Autumn second booster campaign among population aged ‘18+’ or ‘60+’; Vaccine effectiveness is ‘optimistic’(effectiveness as of a booster vaccine against Delta) or ‘pessimistic’ (as against BA.4/BA.5/BA.2.75)

### Entire projection period

*Projections over June 2022 through June 2023*

#### Death

A. Size and timing of peaks. Boxplots show summary of the likely value at peak incidence (median and interquartile range); points show timing and size of peaks from independent sample simulations

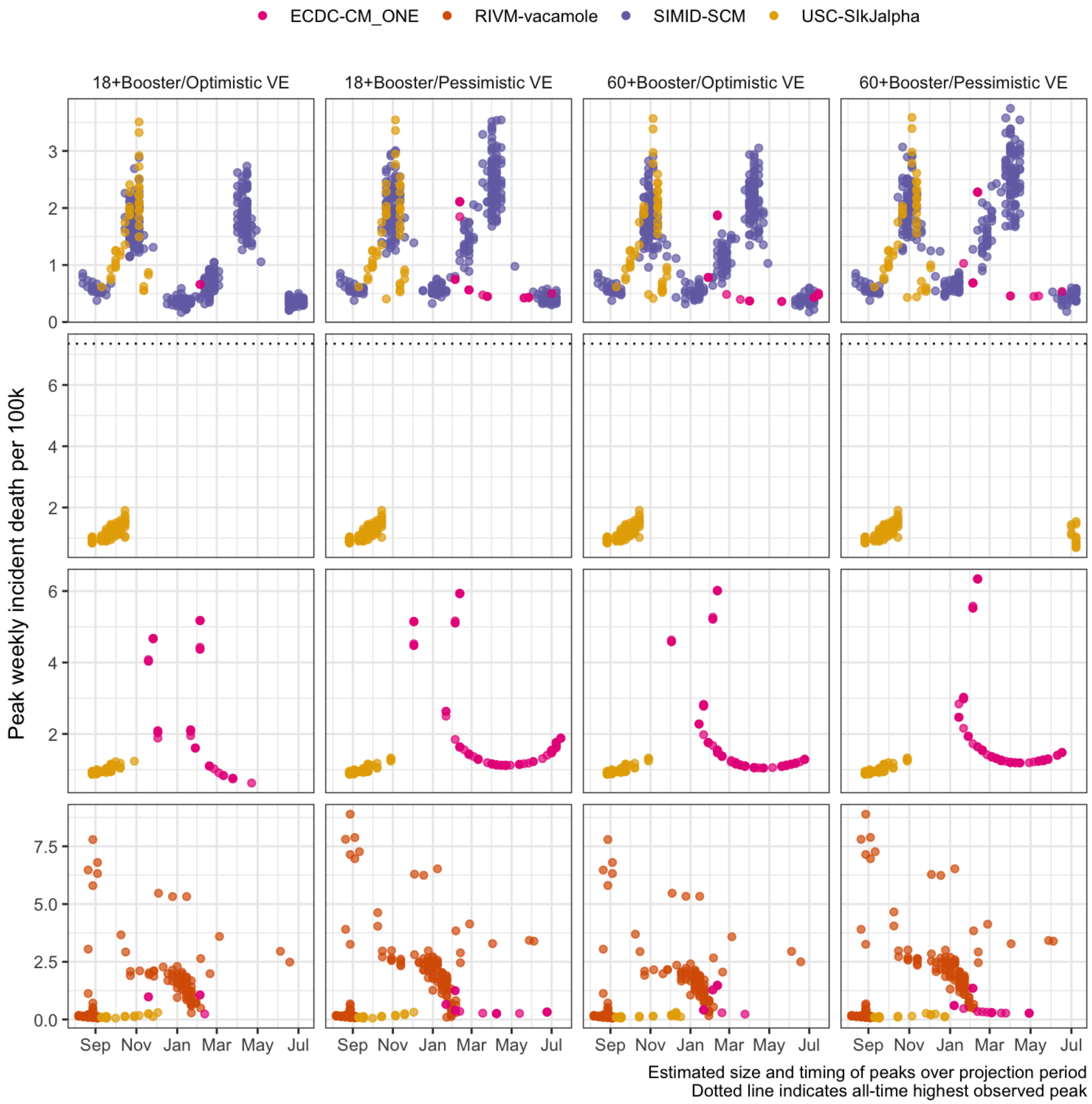


B. Projected number of peaks (median with 5-95% probability)

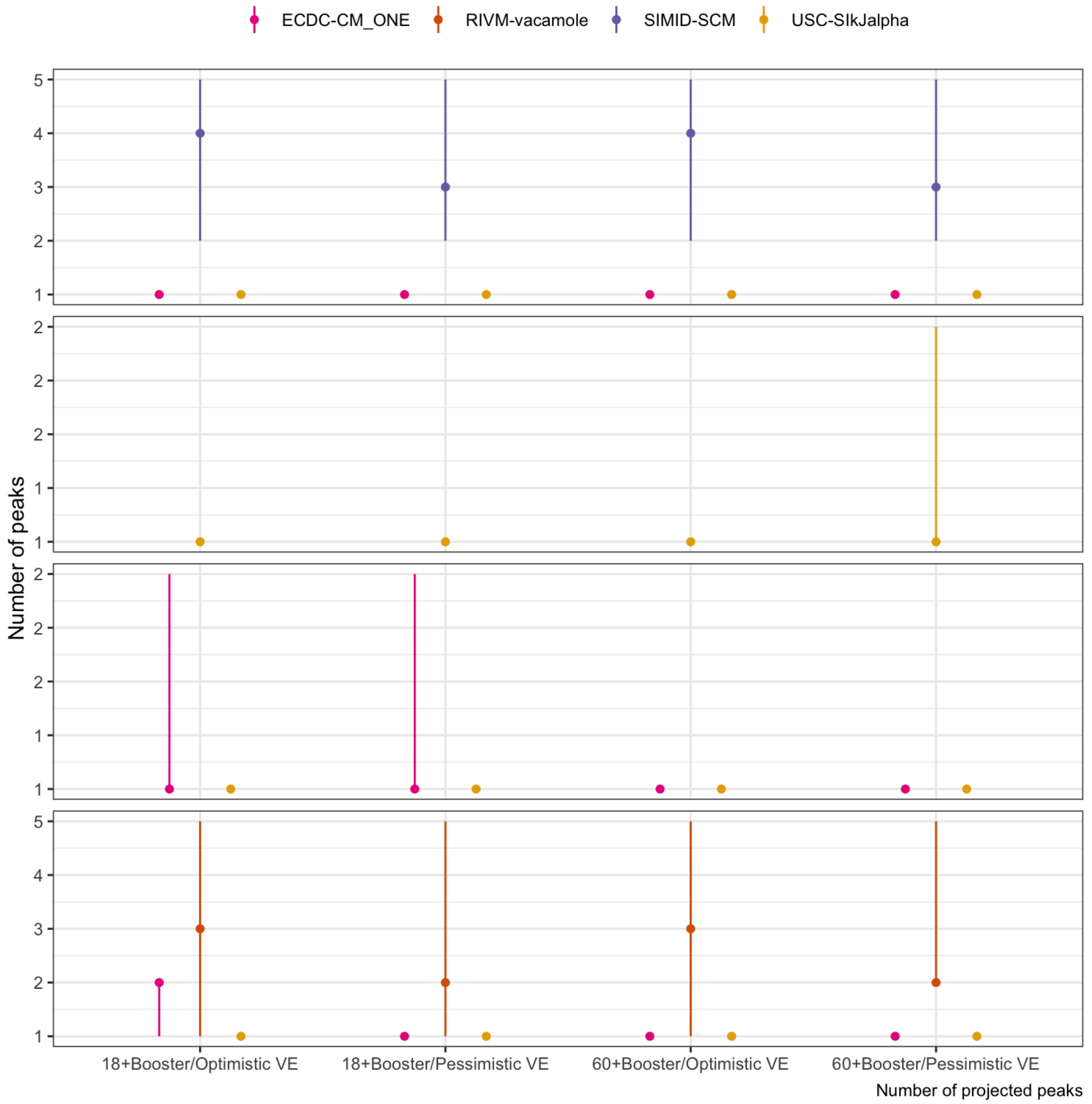


Scenarios: Autumn second booster campaign among population aged ‘18+’ or ‘60+’; Vaccine effectiveness is ‘optimistic’(effectiveness as of a booster vaccine against Delta) or ‘pessimistic’ (as against BA.4/BA.5/BA.2.75)

Case

A. Size and timing of peaks. Boxplots show summary of the likely value at peak incidence (median and interquartile range); points show timing and size of peaks from independent sample simulations

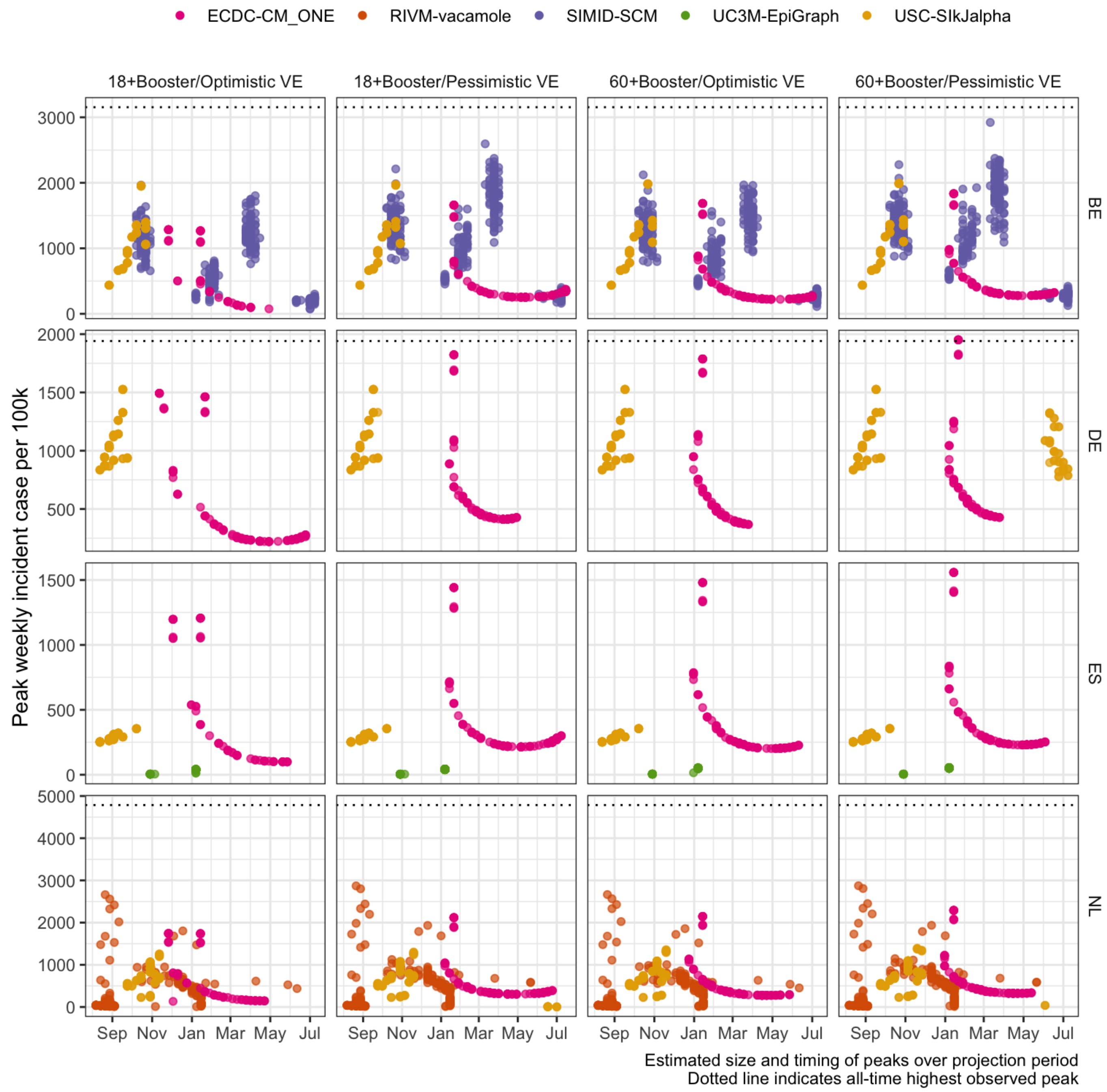


B. Projected number of peaks (median with 5-95% probability)

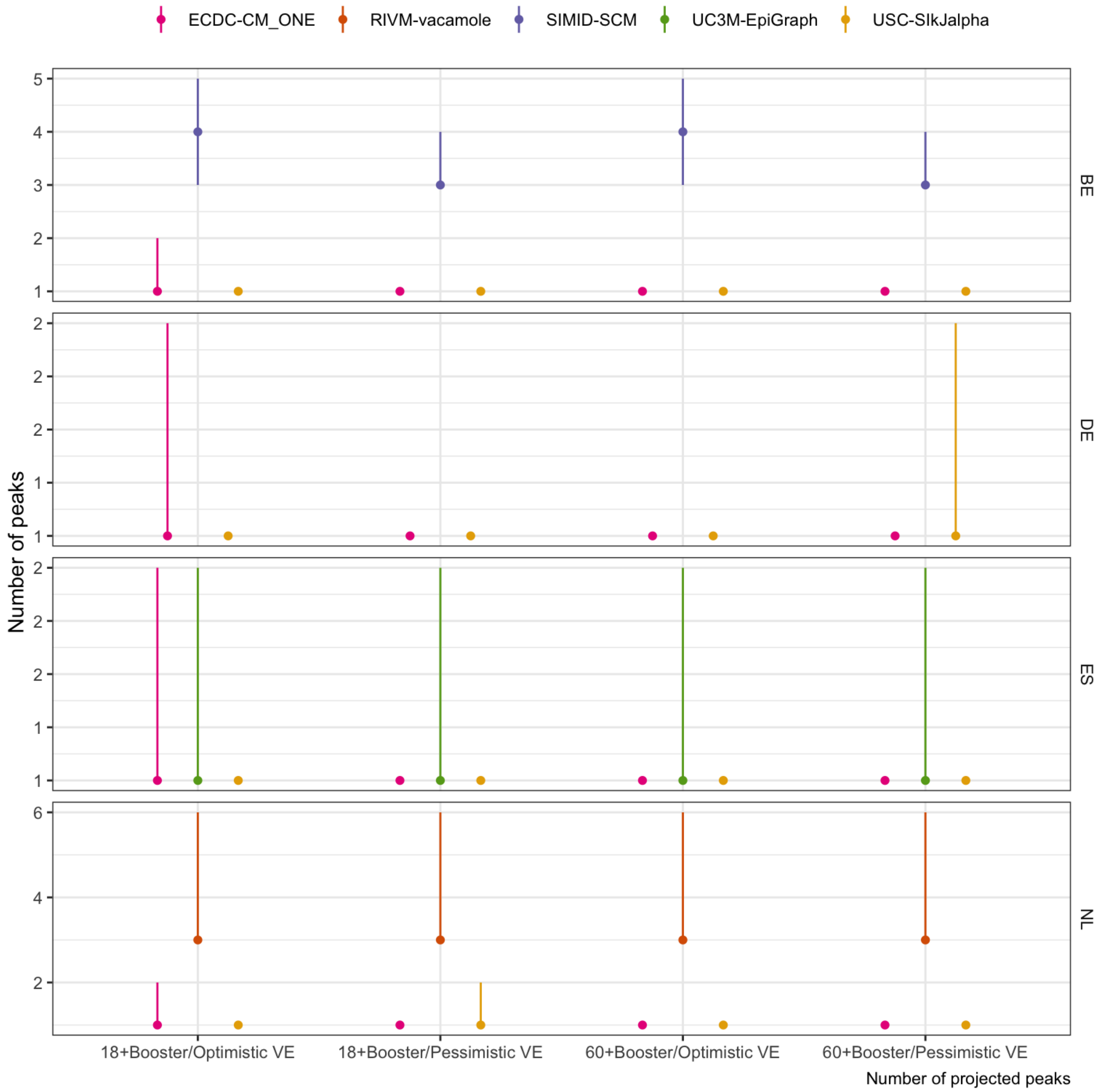


Scenarios: Autumn second booster campaign among population aged ‘18+’ or ‘60+’; Vaccine effectiveness is ‘optimistic’(effectiveness as of a booster vaccine against Delta) or ‘pessimistic’ (as against BA.4/BA.5/BA.2.75)

#### Infection

A. Size and timing of peaks. Boxplots show summary of the likely value at peak incidence (median and interquartile range); points show timing and size of peaks from independent sample simulations

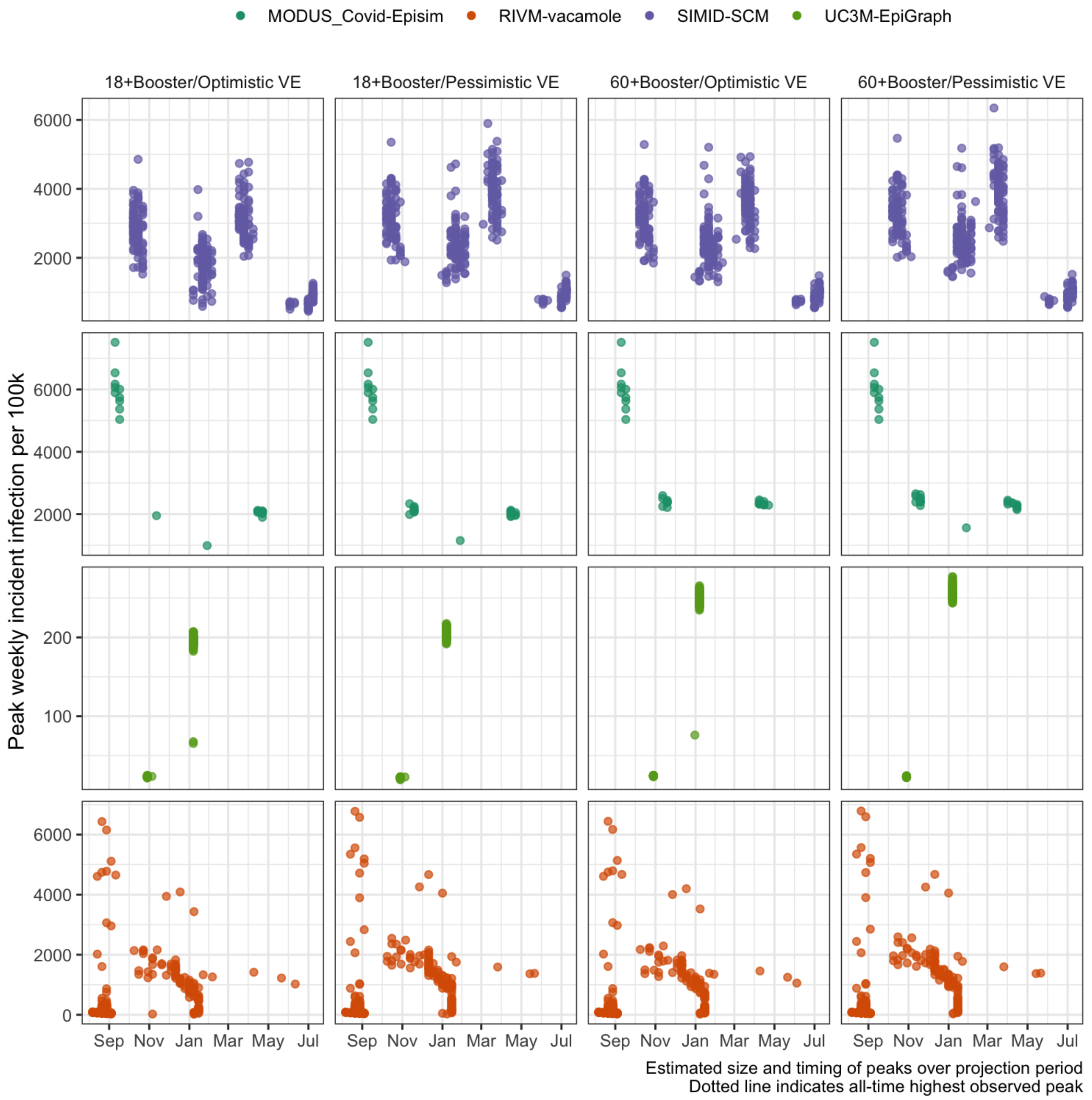


B. Projected number of peaks (median with 5-95% probability)

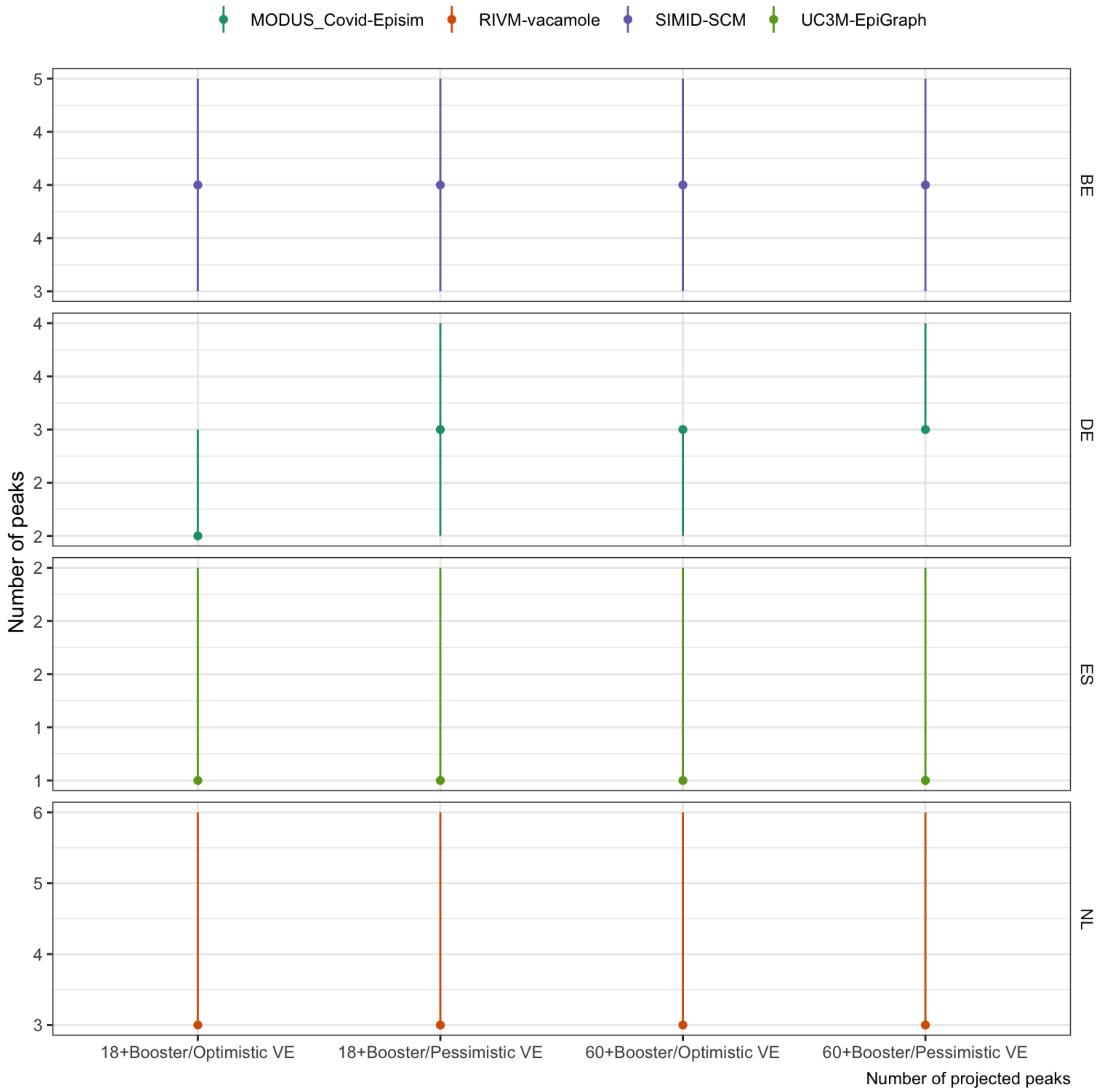


Scenarios: Autumn second booster campaign among population aged ‘18+’ or ‘60+’; Vaccine effectiveness is ‘optimistic’(effectiveness as of a booster vaccine against Delta) or ‘pessimistic’ (as against BA.4/BA.5/BA.2.75)

#### Hosp

A. Size and timing of peaks. Boxplots show summary of the likely value at peak incidence (median and interquartile range); points show timing and size of peaks from independent sample simulations

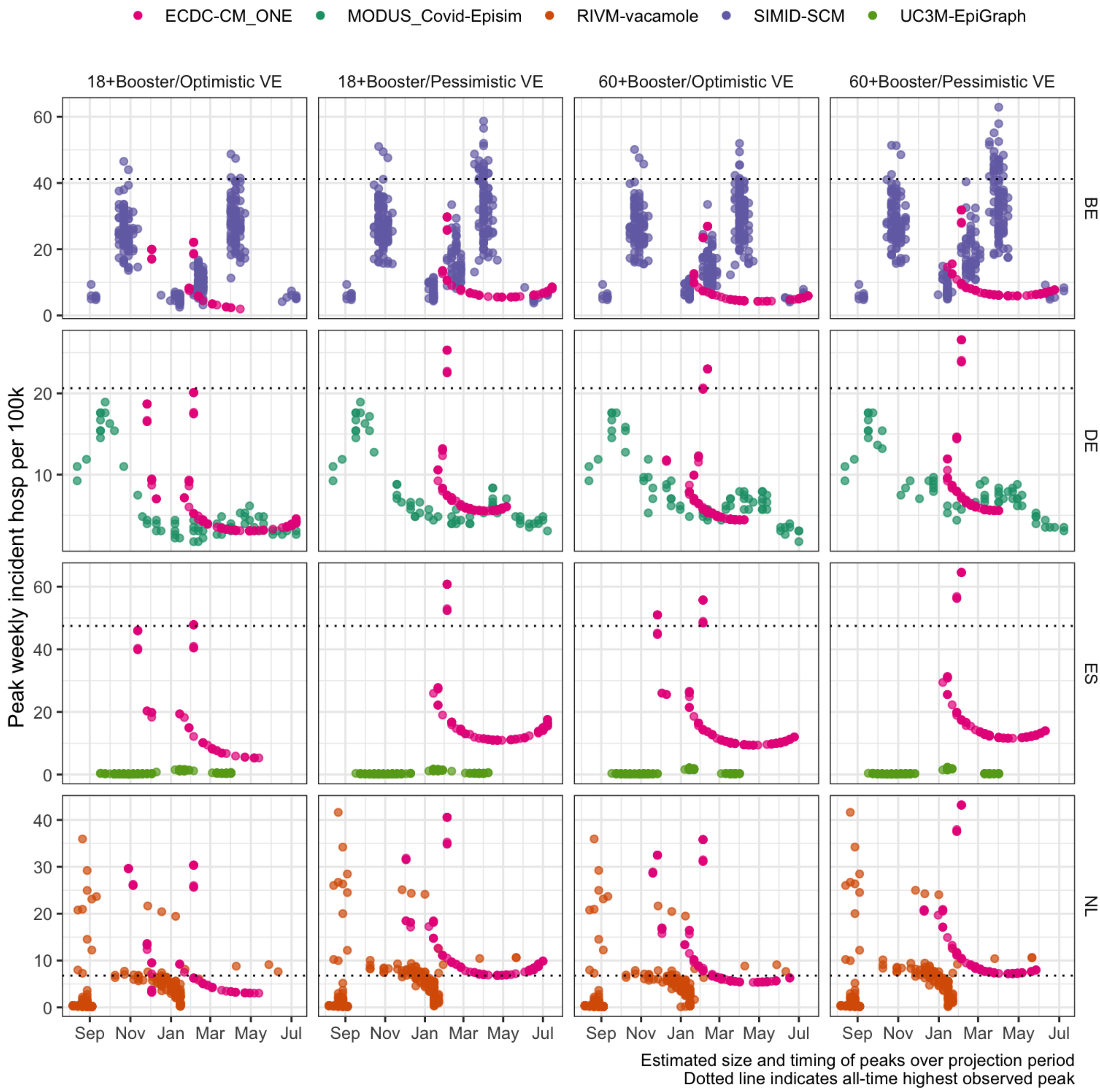


B. Projected number of peaks (median with 5-95% probability)

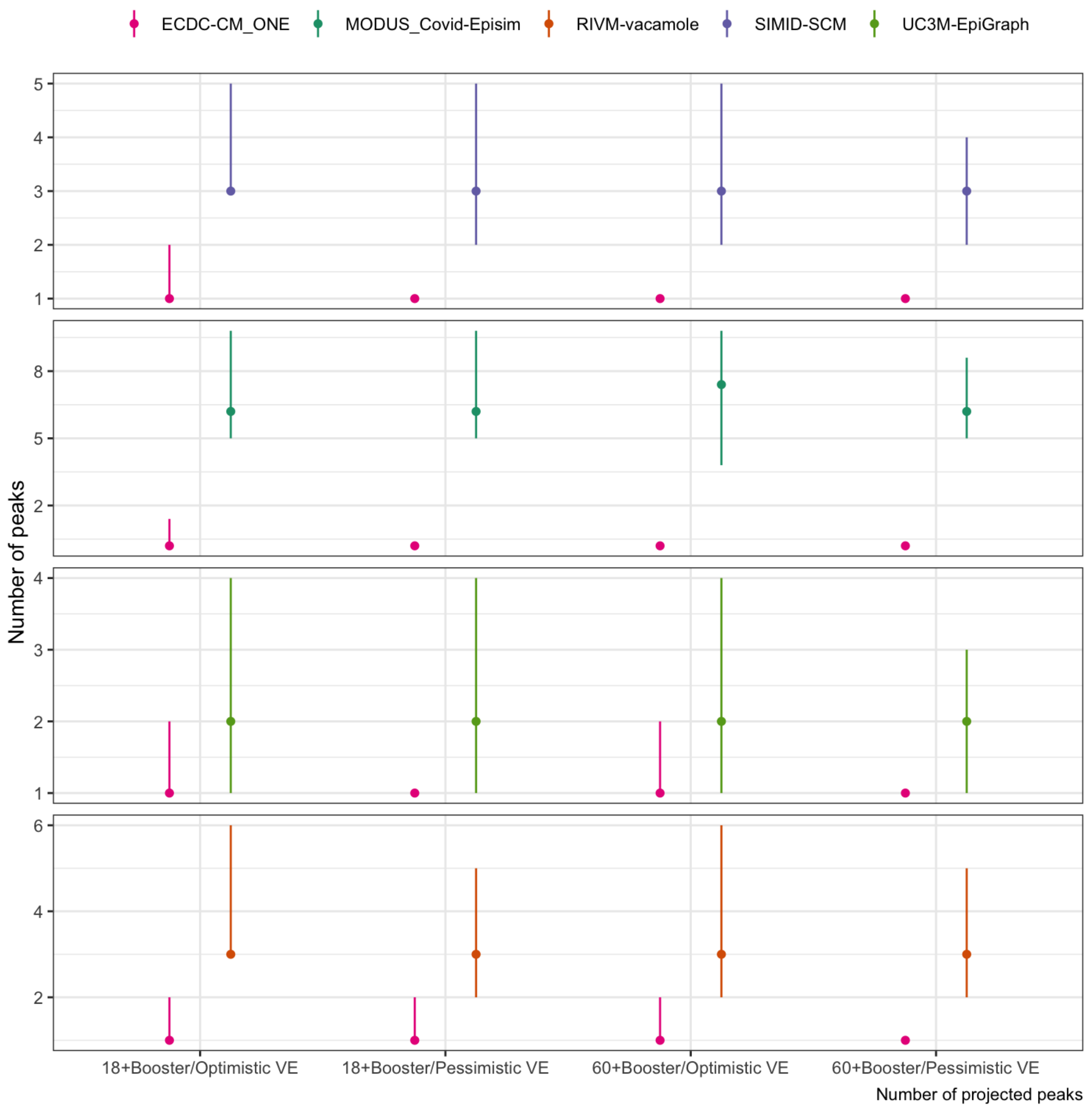


Scenarios: Autumn second booster campaign among population aged ‘18+’ or ‘60+’; Vaccine effectiveness is ‘optimistic’(effectiveness as of a booster vaccine against Delta) or ‘pessimistic’ (as against BA.4/BA.5/BA.2.75)

Icu

A. Size and timing of peaks. Boxplots show summary of the likely value at peak incidence (median and interquartile range); points show timing and size of peaks from independent sample simulations

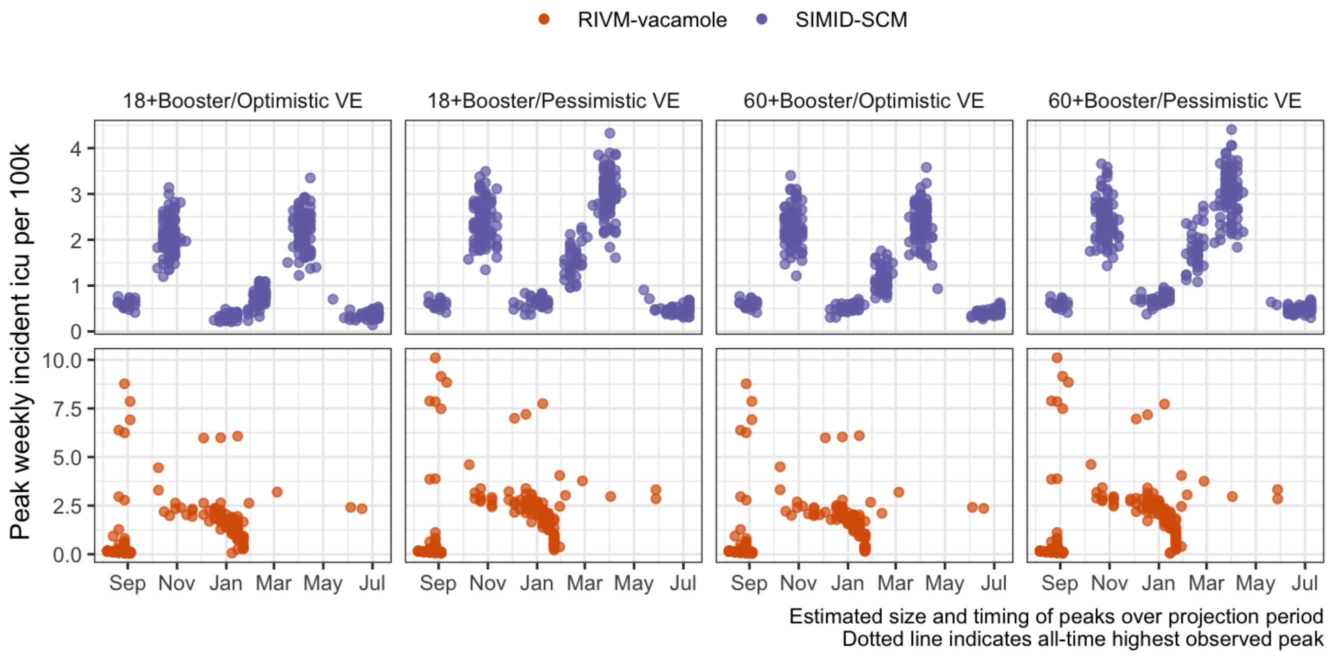


B. Projected number of peaks (median with 5-95% probability)

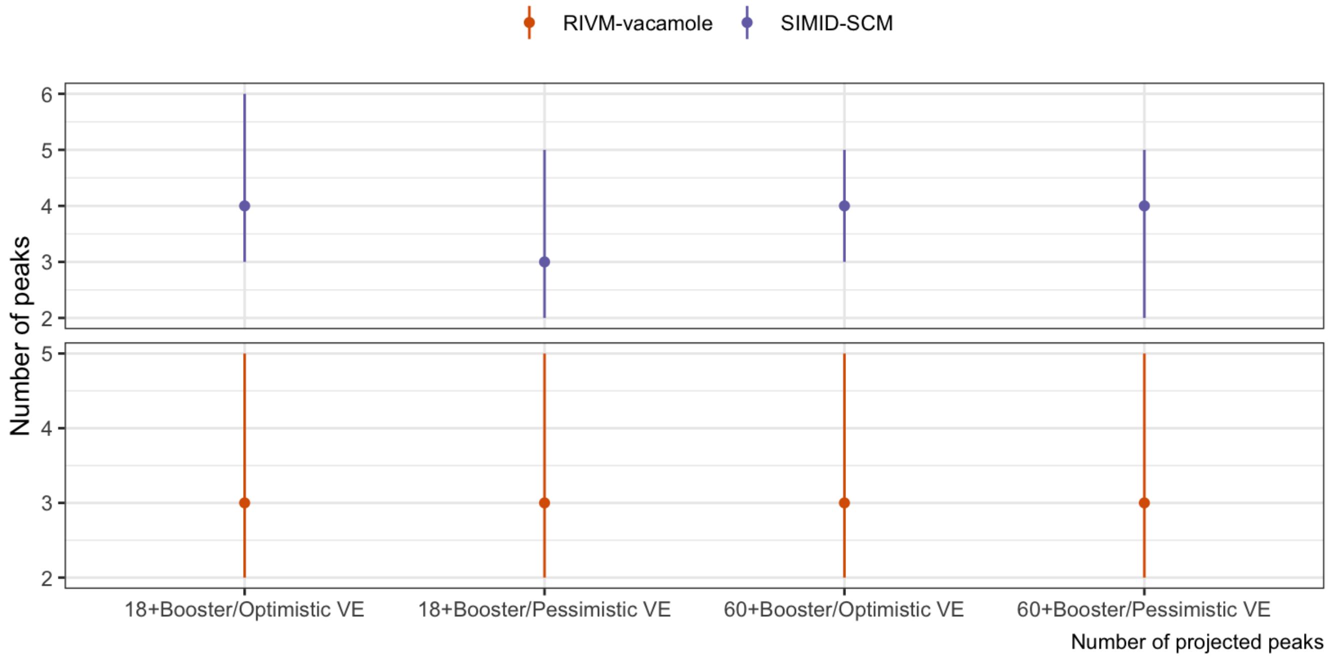


Scenarios: Autumn second booster campaign among population aged ‘18+’ or ‘60+’; Vaccine effectiveness is ‘optimistic’(effectiveness as of a booster vaccine against Delta) or ‘pessimistic’ (as against BA.4/BA.5/BA.2.75)

---

The European Scenario and Forecast (https://covid19forecasthub.eu/index.html) Hubs are run in collaboration between the Epiforecasts team (https://epiforecasts.io/) at the London School of Hygiene & Tropical Medicine (https://www.lshtm.ac.uk); and the European Centre for Disease Control and Prevention (ECDC) (https://ecdc.europa.eu).

**Contact us (contact.html)**
